# Effect of Polygenic Scores on the Relationship Between Psychosis and Cognitive Performance

**DOI:** 10.1101/2024.09.10.24313194

**Authors:** Lauren Varney, Krisztina Jedlovszky, Baihan Wang, Stephen Murtough, Marius Cotic, Alvin Richards-Belle, Noushin Saadullah Khani, Robin Lau, Rosemary Abidoph, Andrew McQuillin, Johan Thygesen, Psychosis Endophenotypes International Consortium (PEIC), Genetic Risk and Outcome of Psychosis (GROUP) Investigators, Behrooz Z. Alizadeh, Stephan Bender, Benedicto Crespo-Facorro, Jeremy Hall, Conrad Iyegbe, Eugenia Kravariti, Stephen M Lawrie, Ignacio Mata, Colm McDonald, Robin M Murray, Diana P Prata, Timothea Toulopoulou, Neeltje van Haren, Elvira Bramon

**Affiliations:** Mental Health Neuroscience, Division of Psychiatry, University College London, UK; Division of Biosciences, University College London, UK; Nuffield Department of Population Health, University of Oxford, UK; Department of Genetics and Genomic Medicine, UCL Great Ormond Street Institute of Child Health, University College London, London, UK; Epidemiology and Applied Clinical Research Department, Division of Psychiatry, University College London, London, UK; Department of Psychology, Institute of Psychiatry, Psychology & Neuroscience, King’s College London, UK; Institute of Health Informatics, Faculty of Population Health Sciences, University College London, UK; Department of Epidemiology, University Medical Center Groningen, University of Groningen, Groningen, The Netherlands; Department of Psychiatry, University Medical Center Groningen, University of Groningen, Groningen, The Netherlands; University of Cologne, Faculty of Medicine and University Hospital Cologne, Department of Child and Adolescent Psychiatry, Psychosomatic Medicine and Psychotherapy; Centre for Biomedical Research in the Mental Health Network (CIBERSAM); Hospital Universitario Virgen del Rocío, Instituto de Biomedicina de Sevilla (IBIS)-CSIC, Spain; Department of Psychiatry, University of Sevilla, Sevilla, Spain; Division of Psychological Medicine and Clinical Neurosciences, Cardiff University, UK; Neuroscience and Mental Health Innovation Institute, Cardiff University, UK; Department of Genetics and Genomic Sciences, Icahn School of Medicine at Mount Sinai Hospital, New York, USA; Department of Psychosis Studies, Institute of Psychiatry, Psychology and Neuroscience, King’s College London, London, UK; Division of Psychiatry, University of Edinburgh, Edinburgh, UK; Fundación Argibide, Pamplona (Navarra), Spain; Clinical Science Institute, University of Galway, Galway, Ireland; Centre for Neuroimaging & Cognitive Genomics (NICOG), Clinical Neuroimaging Laboratory, Galway Neuroscience Centre, College of Medicine Nursing and Health Sciences, University of Galway, H91 TK33 Galway, Ireland; Institute of Psychiatry, Psychology and Neuroscience, King’s College London, UK; Instituto de Biofísica e Engenharia Biomédica, Faculdade de Ciências da Universidade de Lisboa, Portugal; Department of Psychology & National Magnetic Resonance Research Center (UMRAM), Aysel Sabuncu Brain Research Centre (ASBAM), Bilkent University, Ankara, Turkey; Department of Psychiatry, Faculty of Medicine, National and Kapodistrian University of Athens, Athens, Greece; Department of Psychiatry, Icahn School of Medicine at Mount Sinai, New York, USA; Department of Child and Adolescent Psychiatry/Psychology, Erasmus Medical Centre, Rotterdam, Netherlands; Camden and Islington NHS Foundation Trust, London, UK

**Keywords:** psychosis, schizophrenia, bipolar disorder, cognition, polygenic score, endophenotype

## Abstract

**Background:** Up to 80% of psychosis patients experience cognitive impairment. High heritability of both psychosis and cognition means cognitive performance could be an endophenotype for psychosis.

**Methods:** Using samples of adults (N=4,506) and children (N=10,981), we investigated the effect of polygenic scores (PGSs) for schizophrenia and bipolar disorder on cognitive performance, and PGSs for intelligence and educational attainment on psychosis symptoms.

**Results:** Schizophrenia PGS was negatively associated with visuospatial processing/problem-solving in the adult sample (beta: −0.0569; 95% confidence interval [CI]: −0.0926, −0.0212) and working memory (beta: −0.0432; 95% CI: −0.0697, −0.0168), processing speed (b: −0.0491; 95% CI: −0.0760, −0.0223), episodic memory (betas: −0.0581 to −0.0430; 95% CIs: −0.0847 to −0.0162), executive functioning (beta: −0.0423; 95% CI: −0.0692, −0.0155), fluid intelligence (beta: −0.0583; 95% CI: −0.0847, −0.0320), and total intelligence (beta: −0.0458; 95% CI: −0.0709, −0.0206) in the child sample. Bipolar disorder PGS was not associated with any cognitive endophenotypes studied. Lower values on the PGS for intelligence were associated with higher odds of psychosis in adults (odds ratio [OR]: 0.886; 95% CI: 0.811– 0.968) and psychotic-like experiences in children (OR: 0.829; 95% CI: 0.777–0.884). In children, a lower polygenic score for educational attainment was associated with greater odds of psychotic-like experiences (OR: 0.771; 95% CI: 0.724–0.821).

**Conclusions:** The relationship between psychosis and cognitive impairment can be demonstrated bidirectionally at the neurobiological level. The effect of schizophrenia PGS on cognitive performance differs across the lifespan and cognitive domains. Specific cognitive domains may therefore be better endophenotypes than overall cognition.

## 1. Introduction

Heritability estimates for both schizophrenia and bipolar disorder are high (79% and 75%, respectively) and there is a large overlap in the genetic variants associated with these disorders (1, 2). Often combined under the broader term “psychosis”, these disorders are characterised by a loss of contact with reality and present with symptoms such as hallucinations and delusions (3). Up to 80% of people with psychosis also experience clinically significant cognitive impairment (4), both in overall cognitive functioning (5) and within specific cognitive domains (6, 7). Deficits are also seen in their unaffected relatives (8–11) and individuals at clinical high-risk of psychosis (12, 13)

The heritability estimate of cognitive ability is around 50% (14), which is similar in both nonpsychiatric populations and populations of people with psychosis (15, 16). Cognitive function has therefore been suggested as an endophenotype for psychosis; a way to bridge the gap between genetics and symptom presentation, to help understand the disease mechanisms (17–20).

Polygenic scores (PGSs) sum up genetic risk for a phenotype based on common variants, and are one way to research endophenotypes (21). Meta-analytic evidence suggests that schizophrenia PGS is significantly associated with overall cognitive performance within the general population but not in those with psychosis (22), possibly because the effect is already captured by diagnosis or simply due to smaller sample sizes (23, 24). Also key is that this association between schizophrenia PGS and cognitive performance appears to be stable over time in those with psychosis, their healthy relatives, and healthy controls (25). However, evidence is mixed on the exact components of cognitive ability affected by schizophrenia PGS (26). The strongest evidence is for a negative effect on performance IQ, attention, and premorbid intelligence (22), while verbal memory, crystalised intelligence, category fluency, and educational attainment appear to be less influenced by genetic liability (27–30).

As for bipolar disorder PGS, the literature is more mixed. Mistry et al. (31) found a negative association between genetic risk for bipolar disorder and executive functioning in childhood. Performance IQ and processing speed were also negatively associated with common genetic components associated with both bipolar disorder and schizophrenia. However, other research has produced little evidence of an association (23, 27, 32, 33), and associations in the opposite direction (34) between bipolar disorder PGS and cognitive performance.

Polygenic scores for cognitive functioning are positively associated with cognitive performance in population samples (23, 35, 36), samples of psychosis cases (23, 35–37), and ultra-high-risk individuals (38), suggesting cognition is influenced by similar genetic mechanisms irrespective of psychosis risk (23). PGSs for childhood intelligence (39), performance IQ (40), and general cognitive ability (41) have all been found to be significantly lower in individuals with psychosis.

The majority of research on psychosis and cognition has been conducted with adult samples. However, schizophrenia is a neurodevelopmental disorder (42, 43), cognitive impairments are already seen in the prodromal phase of psychosis (44–46), and the transition to psychosis is not associated with further cognitive decline (47). Extending this research beyond adult population adds depth to our understanding of the genetic mechanisms behind psychosis. Psychotic-like experiences in childhood have been associated with poorer cognitive functioning (48, 49) and polygenic scores for cognitive performance and educational attainment have each been negatively associated with childhood psychotic-like experiences (50).

### 1.1. Present Study

The aim of the present study was to examine the effect of polygenic scores for schizophrenia and bipolar disorder on performance within a range of cognitive domains in both adults and children. We also examined whether polygenic scores for cognitive performance and educational attainment were associated with psychosis presentation.

## 2. Method and Materials

### 2.1. Participants

#### 2.1.1. Psychosis Endophenotypes International Consortium (PEIC)

The PEIC is a collaborative effort from multiple sites across Europe (UK, the Netherlands, Spain, Germany) and Australia, comprising data from individuals with a diagnosis of psychosis (bipolar disorder, schizophrenia, or other psychotic disorder; hereafter patients), their unaffected relatives, and healthy control participants. All were of European ancestry (33, 51). Relatives and controls were not excluded if they had a personal history of non-psychotic disorder, as long as they were off psychotropic medication for at least 12 months before assessment. Exclusion criteria (for all clinical groups) included a history of neurological disease or previous loss of consciousness due to head injury (11).

#### 2.1.2. Adolescent Brain Cognition Development (ABCD) Study

The ABCD Study^®^ is a longitudinal study from the USA that aims to investigate the impact of various factors on brain development and health/social outcomes. A population-representative sample of children aged 9-10 years was recruited (52). Participants were excluded if they were not fluent in English, had a history of traumatic brain injury, or a current diagnosis of moderate/severe autism spectrum disorder, schizophrenia, intellectual disability, or substance use disorders (53). Data were taken from baseline assessments.

### 2.2. Genotyping, Quality Control, and Imputation

#### 2.2.1. PEIC

DNA was extracted from blood samples of 6,935 participants and sent to the Wellcome Trust Sanger Institute (Cambridge, UK) for initial processing and quality control (QC). After imputation, 6,215,801 SNPs and 4,835 participants remained. Further details available in Bramon et al. (51) and Supplementary Material.

#### 2.2.2. ABCD Study***^®^***

DNA was extracted from blood/saliva samples of 11,880 participants at the Rutgers University Cell and DNA Repository (RUCDR; New Jersey, USA). After post-imputation QC, 11,229,083 SNPs and 11,017 participants remained. Further detail available in Uban et al. (54), Wang et al. (53) and Supplementary Material, as well at https://nda.nih.gov/study.html?id=901.

### 2.3. Relationship Inference and Principal Component Analysis

#### 2.3.1. PEIC and ABCD Study***^®^***

The GENESIS R/Bioconductor package (55, 56) was used to account for familial relatedness and population structure. An unadjusted kinship matrix was generated first using KING-robust 2.2.5 (57) to infer the relatedness of each pair of participants. The SNPRelate package in R 4.0.2 (58) was used to analyse the genotyped data alongside this kinship matrix to estimate ancestrally representative principal components (PCs). An adjusted kinship matrix was then generated to account for these PCs, allowing for an estimation of familial relatedness independent of ancestry. Further details available in Supplementary Material and Wang et al. (53).

### 2.4. Polygenic Score Generation

All polygenic scores were generated using PRS-CSx (59). Reference panels from the 1000 Genomes Project (60) that best matched the ancestries present in the original GWAS (see Supplementary Material). Scores were standardised against the sample mean (in the PEIC sample, the control group mean).

For schizophrenia and bipolar disorder PGSs, summary statistics from the latest Psychiatric Genomic Consortium (PGC) analyses were used (61, 62). As the PEIC data was used in the PGC discovery sample, summary statistics were obtained that excluded this sample to avoid overlap. For educational attainment PGS, summary statistics from the Lee et al. (63) paper were used. For the intelligence PGS, summary statistics from the Savage et al. (64) paper were used.

### 2.5. Cognitive Tests

#### 2.5.1. PEIC

Three tests were administered: block design and digit span from the Wechsler Adult Intelligence Scale, revised version (WAIS-R; (65) or third edition (WAIS-III; (66)), and the Rey Auditory Verbal Learning Test (RAVLT; (67)).

#### 2.5.2. ABCD Study***^®^***

Participants completed 11 neurocognitive tests, seven from the National Institutes of Health (NIH) Toolbox^®^ (Picture Vocabulary; Oral Reading Recognition; Pattern Comparison; List Sorting; Picture Sequence; Flanker; Dimensional Change Card Sort) and four additional tests (RAVLT; Cash Choice Task; Little Man Task; Matrix Reasoning; (68)).

The NIH Toolbox^®^ tests create three composite scores: Crystalised Intelligence, Fluid Intelligence, and Total Intelligence (68). The cognitive domains that each test measures are presented in Table 1 (68–70). Further detail is available in Supplementary Material.

**Table 1.** Cognitive Tests Administered to Participants in the PEIC and ABCD Study^®^ Datasets and the Associated Cognitive Domains.

| Dataset | Cognitive Test | Cognitive Domain |
| --- | --- | --- |
| PEIC | Block Design | Visuospatial processing<br>Non-verbal problem solving |
|  | Digit Span | Working memory<br>Attention |
| PEIC & ABCD Study<sup>®</sup> | Rey Auditory Verbal Learning Task (RAVLT) | Verbal learning<br>Episodic memory<br>Verbal memory |
| ABCD Study<sup>®</sup> | NIH Toolbox Picture Vocabulary Test<sup>®</sup><br>(Crystallised Intelligence Composite) | Language<br>Verbal intellect<br>Verbal comprehension |
|  | NIH Toolbox Oral Reading Recognition Test<sup>®</sup><br>(Crystallised Intelligence Composite) | Language<br>Reading ability<br>Reading decoding<br>Academic achievement |
|  | NIH Toolbox Pattern Comparison Processing Speed Test<sup>®</sup><br>(Fluid Intelligence Composite) | Processing speed<br>Information processing |
|  | NIH Toolbox List Sorting Working Memory Test<sup>®</sup><br>(Fluid Intelligence Composite) | Working memory<br>Categorisation<br>Information processing |
|  | NIH Toolbox Picture Sequence Memory Test<sup>®</sup><br>(Fluid Intelligence Composite) | Visuospatial sequencing<br>Visuospatial memory |
|  | NIH Toolbox Flanker Task<sup>®</sup><br>(Fluid Intelligence Composite) | Cognitive control<br>Attention |
|  | NIH Toolbox Dimensional Change Card Sort<sup>®</sup><br>(Fluid Intelligence Composite) | Flexible thinking<br>Concept formation<br>Set shifting |
|  | Cash Choice Task | Delay of gratification<br>Motivation<br>Impulsivity |
|  | Little Man Task | Visuospatial attention<br>Perspective-taking<br>Mental rotation |
|  | Matrix Reasoning | Visuospatial ability<br>Fluid reasoning<br>Part-whole reasoning<br>Visual sequencing |

*Abbreviations:* PEIC = Psychosis Endophenotypes International Consortium; ABCD = Adolescent Brain Cognitive Development; NIH = National Institutes of Health

### 2.6. Psychosis Outcome

#### 2.6.1. PEIC

All participants underwent a structured clinical interview to confirm/rule out the presence of a DSM-IV diagnosis of schizophrenia or another psychotic disorder (33, 51) and determine psychosis outcomes. This led to three clinical groups: patients (n = 1,231), relatives (n = 856), and controls (n = 2,740).

#### 2.6.2. ABCD Study***^®^***

Responses on the Prodromal Questionnaire–Brief Child Version (PQ-BC), a 21-item self-report questionnaire of psychotic-like experiences in the past month, were used to measure psychotic-like presentations. Each item has three parts: whether they experienced the symptom; if yes, whether it was distressing; and, if yes, how distressing on a scale of 1 (“not very bothered”) to 5 (“extremely bothered”). Scores of 3 (“moderately bothered”) or more were classed as significantly distressing (49).

### 2.7. Statistical Analyses

Mixed model regression analyses (linear/logistic) were used to investigate the effect of PGSs on cognitive performance and psychosis presentation. Age, sex, and ancestry PCs were included as fixed effects (as well as research site for all PEIC analyses, and clinical group in the cognitive performance PEIC analyses); the adjusted kinship matrix was included as a random effect. Linear cognitive test scores were standardised against the group mean or control group mean in the ABCD Study^®^ sample and PEIC samples, respectively. Interaction and subgroup analyses were also conducted within the PEIC sample to determine whether the effect of PGS differed between clinical groups.

A multiple testing correction of 0.05/4 (three cognitive tests and group status prediction) was applied to the analyses carried out with the PEIC sample, leaving an adjusted significance threshold of *p* < .0125. In the ABCD Study^®^ dataset, cognitive tests were grouped by domain. This resulted in seven domains (Supplementary Table S1) which, combined with the psychotic-like experience prediction, lead to an adjusted significance threshold of *p* < .00625 (0.05/8). Uncorrected *p*-values are reported but interpreted using the respective adjusted threshold.

We used a Support Vector Machine (SVM) supervised machine learning algorithm to predict the clinical group status (patient, relative, or control) from cognition-related polygenic risk scores and demographic parameters (age, sex, ancestry PCs, research sites), following the method from Bracher Smith et al. (71). This was included as a robust and replicable baseline to validate and benchmark the regression. To assess the relative importance of PGS compared to demographic predictors on clinical group status, the SVM model was further inspected using permutation feature importance (72). Further details of these analyses can be found in the Supplementary Material.

### 2.8. Ethics Statement

#### 2.8.1. PEIC

The Psychosis Endophenotypes International Consortium was approved by the local ethics committee at each participating research centre, and all participants provided written informed consent before assessment.

#### 2.8.2. ABCD Study***^®^***

Informed assent/consent was obtained from participants and their parents at the research centre they were recruited at.

## 3. Results

### 3.1. Demographics

Demographic data for participants in the PEIC sample are presented in Table 2 and for the ABCD Study^®^ sample in Table 3. In both samples, the split between males and females was relatively equal (though in the PEIC sample subgroups, there were more male patients and more female relatives). In the ABCD sample, just over half were of White ethnicity.

**Table 2.** Demographic Characteristics for Participants in the PEIC Sample.

|  |  |  |  |  |
| --- | --- | --- | --- | --- |
|  | **Patients** | **Relatives** | **Controls** | **Total** |
| **Sample size, N** | 1,182 | 854 | 2,470 | 4,506 |
| **Age, mean years (SD)** | 33.5 (10.4) | 45.7 (15.9) | 45.5 (16.2) | 42.4 (15.8) |
| Age range | 15 - 79 | 16 - 85 | 16 - 84 | 15 - 85 |
| **Gender, n (%)** |  |  |  |  |
| Male | 794 (67.2%) | 344<br>(40.3%) | 1,182<br>(47.9%) | 2,320<br>(51.5%) |
| Female | 388 (32.8%) | 510<br>(59.7%) | 1,288<br>(52.1%) | 2,186<br>(48.5%) |
| **Centre, n (%)** |  |  |  |  |
| Edinburgh | 31 (2.6%) | - | 17 (< 0.1%) | 48 (1.1%) |
| Heidelberg | 24 (2.0%) | 8 (0.9%) | 22 (0.9%) | 55 (1.2%) |
| Holland | 370 (31.3%) | 505<br>(59.1%) | 974 (39.4%) | 1,849<br>(41.0%) |
| London | 237 (20.0%) | 197<br>(23.1%) | 324 (13.1%) | 758<br>(16.8%) |
| Munich | - | - | 962 (38.9%) | 962<br>(21.3%) |
| Pamplona | 44 (3.7%) | - | - | 44 (1.0%) |
| Perth | 309 (26.1%) | 143<br>(16.7%) | 163 (6.6%) | 615<br>(13.6%) |
| Santander | 167 (14.1%) | - | 8 (0.3%) | 175 (3.9%) |
| **Diagnosis, n (%)** |  |  |  |  |
| Anxiety and/or<br>Depressive Disorder | - | 192<br>(22.5%) | 173 (7.0%) | 365 (8.1%) |
| Bipolar Disorder | 107 (9.1%) | - | - | 107 (2.4%) |
| Other Psychotic<br>Disorder | 169 (14.3%) | - | - | 169 (3.8%) |
| Personality Disorder | - | 1 (0.1 %) | - | 1 (< 0.1%) |
| Schizophrenia | 906 (76.6%) | - | - | 906<br>(20.1%) |
| Substance Misuse | - | 4 (0.5%) | 11 (0.4%) | 15 (0.3%) |
| No Psychiatric<br>Disorder | - | 657<br>(76.9%) | 2,286<br>(92.6%) | 2,943<br>(65.3%) |
| *Abbreviations: PEIC = Psychosis Endophenotypes International Consortium; N/n = number of participants; SD = standard deviation* |  |  |  |  |

**Table 3.** Baseline Demographic Characteristics for Participants in the ABCD Sample.

|  |  |
| --- | --- |
| **Sample size**, N (%) | 10,981 |
| **Age**, years (mean, SD) | 9.9 (0.63) |
| Age range, years | 8.9 – 11.1 |
| **Gender**, n (%) |  |
| Male | 5,805 (52.9%) |
| Female | 5,176 (47.1%) |
| **Ethnicity**, n (%) |  |
| White | 5,872 (53.5%) |
| Black | 1,684 (15.3%) |
| Hispanic | 2,099 (19.1%) |
| Asian | 155 (1.4%) |
| Other | 1,169 (10.6%) |
| Missing | 2 (< 0.1%) |
| **Psychotic-Like Symptoms (PQ-B)**, n (%) |  |
| At Least One Symptom | 6,735 (61.3%) |
| Distress From At Least One Symptom | 4,748 (43.2%) |
| Significant Distress From At Least One Symptom | 2,931 (26.7%) |
| Missing | 5 (< 0.1%) |
| *Abbreviations:* ABCD = Adolescent Brain Cognitive Development; N/n = number of participants; SD = standard deviation; PQ-B = Prodromal Questionnaire – Brief version |  |

### 3.2. Cognitive Test Performance

Details of average cognitive performance for both the PEIC sample and ABCD sample are available in the Supplementary Material and Supplementary Tables S2-S4. When comparing PEIC subgroups, controls performed significantly better than patients on all tests;

### 3.3. Effect of Polygenic Scores on Cognitive Performance

#### 3.3.1. Effect of Polygenic Scores on Cognitive Performance: PEIC (Adult) Sample

Schizophrenia PGS was negatively associated with block design performance (b: −0.0569; 95% CI: −0.0926, −0.0212; *p* = .00179), but no associations were identified with other cognitive tests (*p*s > .356). Bipolar disorder PGS showed no significant associations after multiple testing correction (*p*s > .0219). Intelligence and educational attainment polygenic scores were each significantly positively associated with performance on all tests (*p*s < .000897) (Figure 1; Supplementary Tables S5 and S6). Interaction analyses between PGS and clinical group were non-significant. Interaction and subgroup analyses are discussed in detail in Supplementary Tables S7 and S8 and the Supplementary Material.

**Figure 1.**
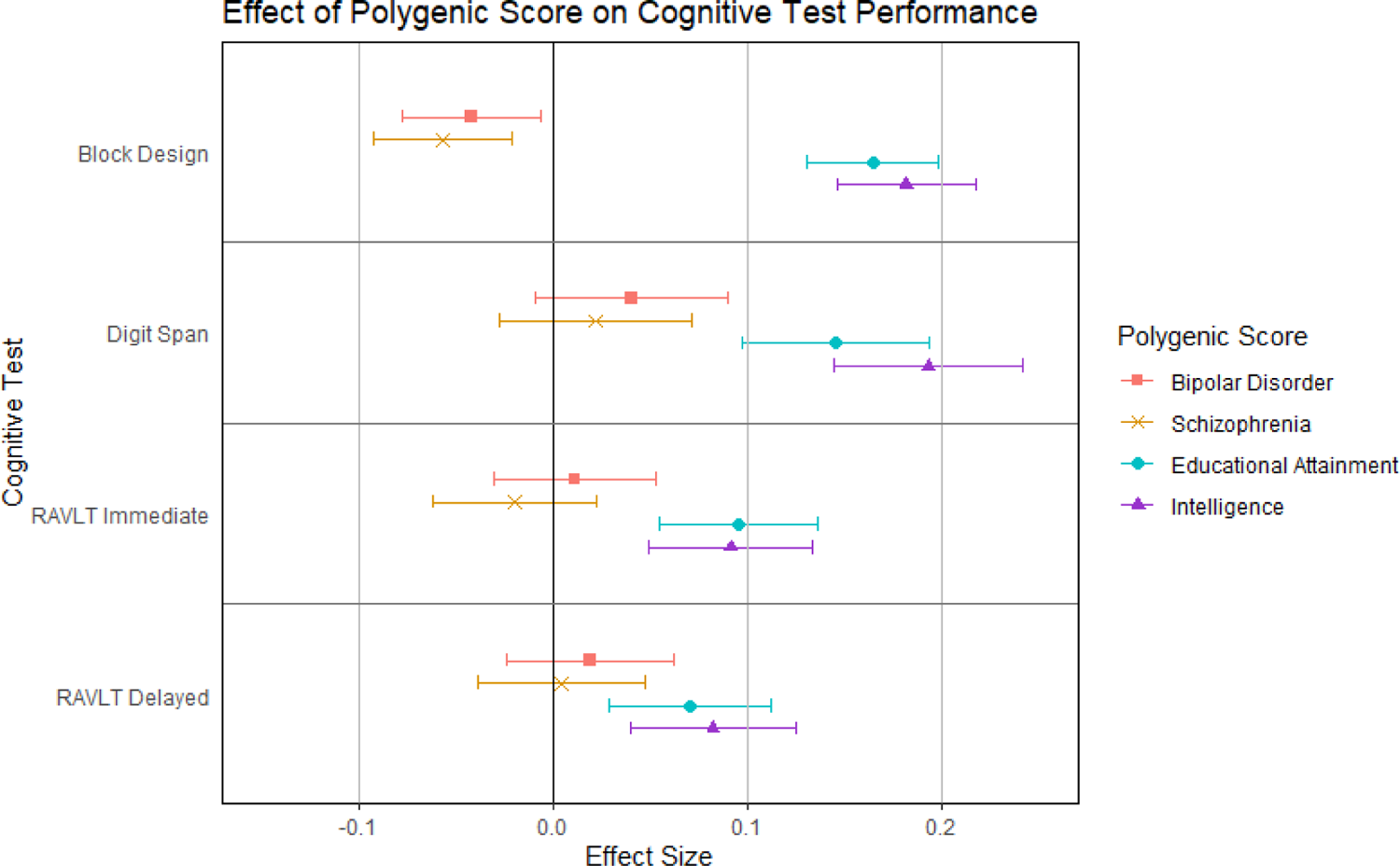
Effect of Polygenic Scores on Cognitive Test Performance in the PEIC Sample. Effect of psychosis-related and cognition-related polygenic scores on cognitive test performance in adults, while controlling for the effect of age, gender, clinical group (patient/relative/control), research site, ancestry (the first four ancestry principal components), and participant inter-relatedness (kinship matrix). Scores have been standardised using the mean and standard deviation from the control group. Standardised values are given in Supplementary Table S5; non-standardised values are given in Supplementary Table S6. *Abbreviations:* PEIC = Psychosis Endophenotype International Consortium; RAVLT = Rey Auditory Verbal Learning Task

#### 3.3.2. Effect of Polygenic Scores on Cognitive Performance: ABCD Study**^®^**(Child) Sample

Higher schizophrenia PGS was associated with poorer performance on: Card Sort (b: −0.0423; 95% CI: −0.0692, −0.0155, *p* = .00201), List Sorting (b: −0.0432; 95% CI: −0.0697, −0.0168; *p* = .00136), Pattern Comparison (b: −0.0491; 95% CI: −0.0760, −0.0223; *p* = .000342), and Picture Sequence (b: −0.0430; 95% CI: −0.0697, −0.0162; *p* = .00164) from the NIH Toolbox^®^, and immediate (b: −0.0437, 95% CI: −0.0699, −0.0174; *p* = .00111), short-delayed (b: −0.0581; 95% CI: −0.0847, −0.0315; *p* = 1.92×10^-5^), and long-delayed (b: −0.0483; 95% CI: −0.0750, −0.0216; *p* = .000397) recall on the RAVLT (Figure 2). Schizophrenia PGS was also negatively associated with Fluid Intelligence (b: −0.0583; 95% CI: −0.0847, −0.0320; *p* = 1.44×10^-5^) and Total Intelligence (b: −0.0458; 95% CI: −0.0709, −0.0206; *p* = .000362). The effect of schizophrenia PGS on Crystalised Intelligence was not significant (*p* = .162). There were no significant effect of bipolar disorder PGS (*p*s > .0185) on cognitive test performance. Intelligence and educational attainment polygenic scores were significantly associated with performance on all tests and composites (*p*s < .00381). Full results are presented in Supplementary Tables S9-S14.

**Figure 2.**
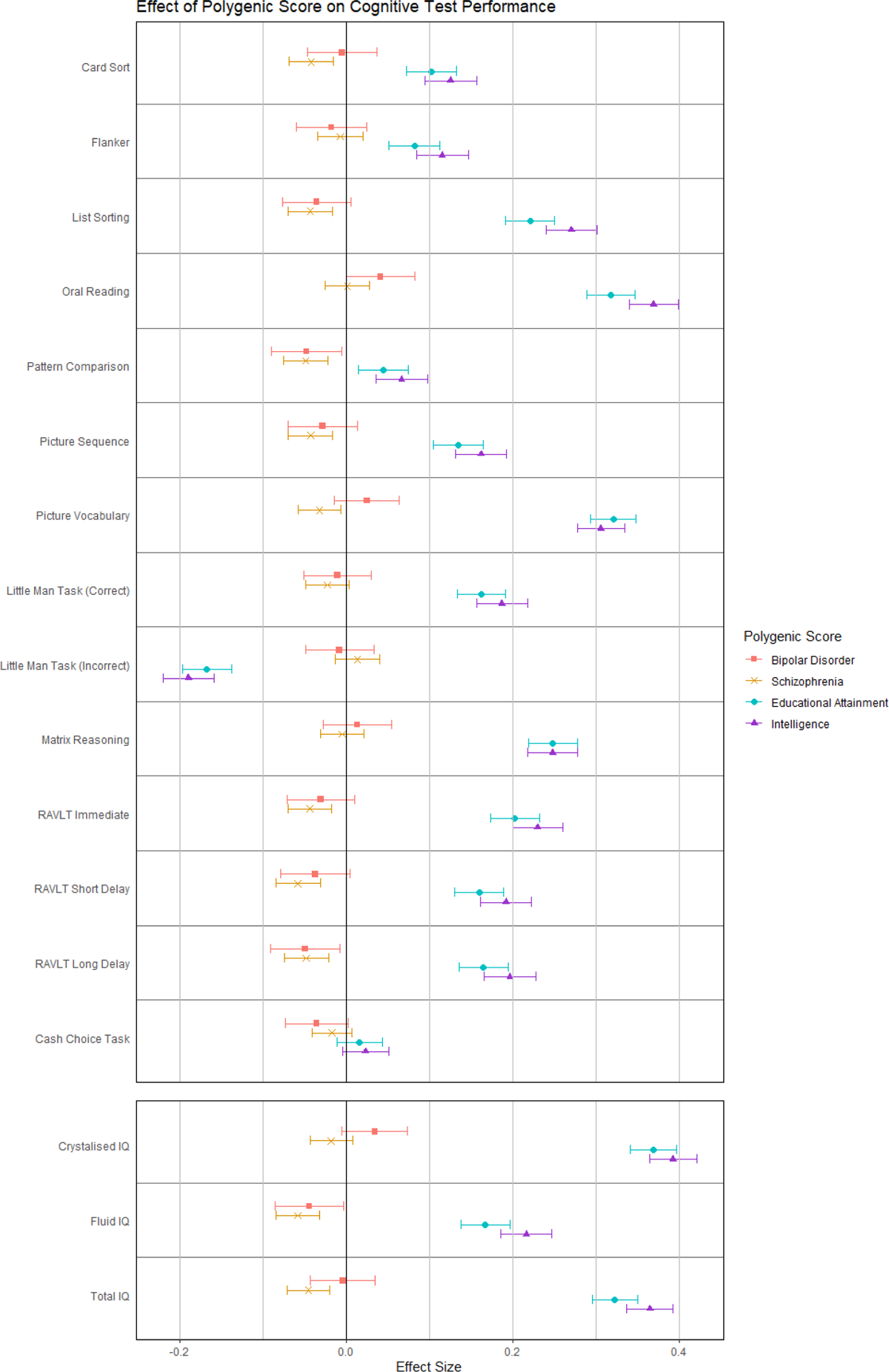
Effect of Polygenic Scores on Performance on Individual Cognitive Tests and Composite Intelligence Scores in the ABCD Study**^®^** Sample Effect of psychosis-related and cognition-related polygenic scores on cognitive test performance in children, while controlling for the effect of age, gender, ancestry (the first eight ancestry principal components), and participant inter-relatedness (kinship matrix). Scores have been standardised using the mean and standard deviation from the whole sample. Standardised values are given in Supplementary Table S9 and S11; non-standardised values are given in Supplementary Table S10 and S12. *Note:* “Little Man Task (Correct)” refers to the percentage of participants’ responses were correct during the task. “Little Man Task (Incorrect)” refers to the percentage of participants’ responses were incorrect. Cash Choice Task effect is the log-transformed result from a logistic regression analysis. Effect > 0 indicates greater odds of choosing the delayed gratification option ($115 in three months); effect < 0 indicates greater odds of choosing the immediate gratification option ($75 in three days). Log-transformed values are given in Supplementary Table S13; logistic regression results are given in Supplementary Table S14. *Abbreviations:* ABCD = Adolescent Brain Cognitive Development; RAVLT = Rey Auditory Verbal Learning Task

### 3.4. Effect of Polygenic Scores on Psychosis Presentation

#### 3.4.1. Effect of Polygenic Scores on Psychosis Presentation: PEIC (Adult) Sample

The logistic regression models showed that intelligence PGS was able to distinguish between patients and controls (OR: 0.886; 95% CI: 0.811–0.968; *p* = .00719) and between relatives and controls (though this latter finding was only a statistical trend after correction for multiple comparisons; *p* = .0190; Figure 3). Educational attainment PGS was a poor predictor of clinical group in the logistic regression analyses (*p*s > .445; Supplementary Table S15). Machine learning analysis validated these findings, as the SVM model including intelligence PGS also distinguished well between the patients and controls (AUROC_median_: 0.847) and between patients and relatives (AUROC_median_: 0.781). The educational attainment PGS model also classified patients from controls (AUROC_median_: 0.856) and patients from relatives (AUROC_median_: 0.752) with high accuracy. However, the polygenic scores themselves did not show a considerable relative importance in any of these analyses. Further details available in the Supplementary Material. Both bipolar disorder PGS (*p*s < 2.38×10^-5^) and schizophrenia PGS (*p*s < 6.80×10^-7^) were able to significantly distinguish between all participant groups.

**Figure 3.**
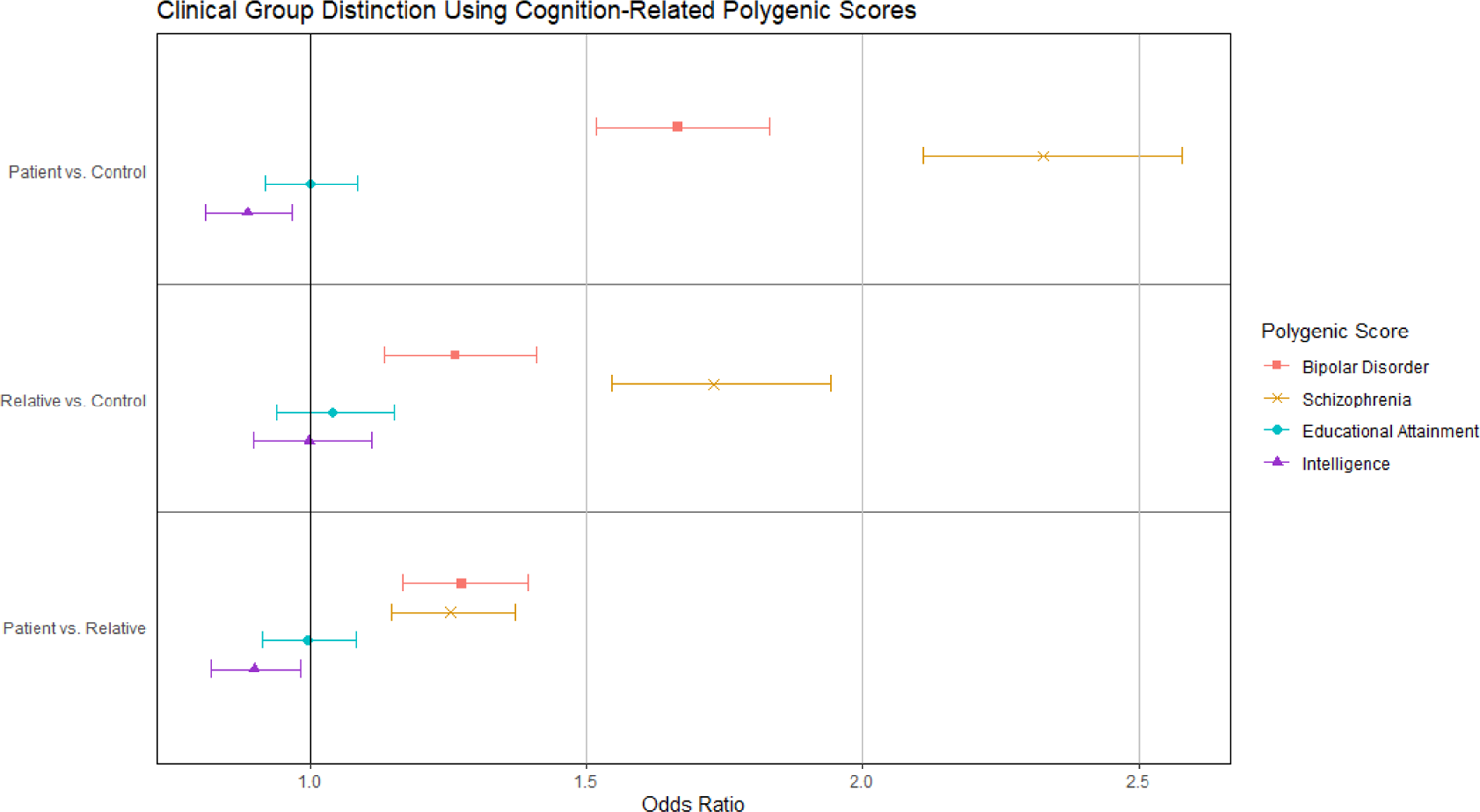
Prediction of Clinical Group Status Using Cognition-Related Polygenic Scores in the PEIC Sample. Effect of psychosis-related and cognition-related polygenic scores on the odds of being in the comparison group (i.e. patient, relative, patient, respectively from top to bottom) compared to the odds of being in the comparison group (i.e. control, control, relative, respectively). Values are given in Supplementary Table S15. *Abbreviations:* PEIC = Psychosis Endophenotypes International Consortium

#### 3.4.2. Effect of Polygenic Scores on Psychosis Presentation: ABCD Study^®^ (Child) Sample

The regression models showed that educational attainment PGS significantly distinguished between children who experienced psychotic-like experiences at baseline and those who did not (OR: 0.771; 95% CI: 0.724–0.821; *p* = 5.86×10^-16^), those who experienced distressing psychotic-like experiences and those who did not (OR: 0.813, 95% CI: 0.764–0.864, *p* = 5.48×10^-11^), and those who experienced significantly distressing psychotic-like experiences and those who did not (OR: 0.769; 95% CI: 0.717–0.826; *p* = 3.87×10^-13^) (Figure 4 and Supplementary Table S18). Educational attainment PGS was also significantly negatively associated with the number of each category of psychotic-like experiences reported (Table 4).

**Figure 4.**
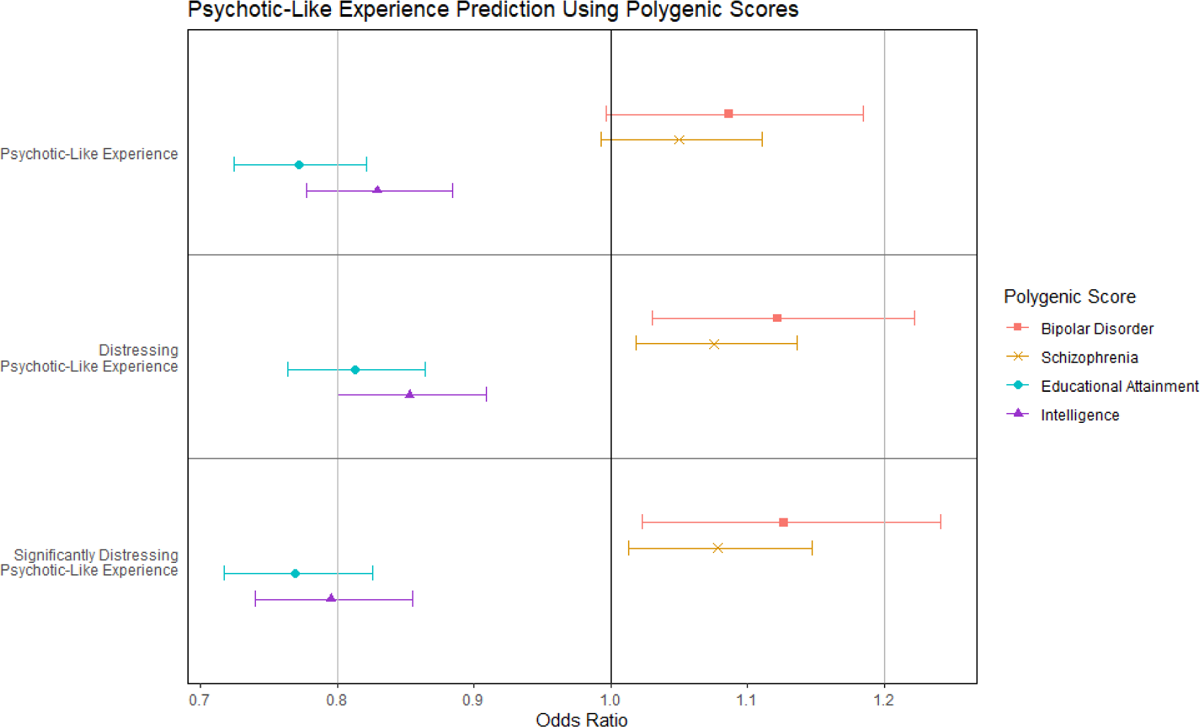
Prediction of Psychotic-Like Experiences at Baseline Using Polygenic Scores in the ABCD Study**^®^**Sample Effect of psychosis-related and cognition-related polygenic scores on the odds of experiencing at least one of the given types of psychotic-like experiences (i.e. odds of experiencing at least one psychotic-like experience, at least one distressing psychotic-like experience, at least one significantly distressing psychotic-like experience, respectively from top to bottom) compared to the odds of having such experiences. Psychotic-like experiences measured using the Prodromal Questionnaire–Brief Child Version (PQ-BC). Values given in Supplementary Table S14. *Abbreviations:* ABCD = Adolescent Brain Cognitive Development

**Table 4.** Effect of Polygenic Scores on Psychotic-Like Experiences at Baseline by Participants in the ABCD Study^®^ Sample.

| **Schizophrenia PGS** | **Mean Difference** | **95% CI** | **R<sup>2</sup> of PGS** | **p** |
| --- | --- | --- | --- | --- |
| Number of psychotic-like experiences | 0.110 | 0.0134, 0.207 | 0.000470 | .0257 |
| Number of distressing psychotic-like experiences | 0.0608 | -0.00176, 0.123 | 0.000342 | .0568 |
| Number of significantly distressing psychotic-like experiences | 0.162 | -0.0410, 0.365 | 0.000230 | .118 |
| **Bipolar Disorder PGS** | **Mean Difference** | **95% CI** | **R<sup>2</sup> of PGS** | **p** |
| Number of psychotic-like experiences | 0.0780 | -0.0721, 0.308 | 9.81x10<sup>-5</sup> | .308 |
| Number of distressing psychotic-like experiences | 0.0600 | -0.0369, 0.157 | 0.000139 | .225 |
| Number of significantly distressing psychotic-like experiences | 0.194 | -0.120, 0.509 | 0.000138 | .226 |
| **Educational Attainment PGS** | **Mean Difference** | **95% CI** | **R<sup>2</sup> of PGS** | **p** |
| Number of psychotic-like experiences | -0.369 | -0.477, -0.261 | 0.00422 | 2.52x10<sup>-11</sup> |
| Number of distressing psychotic-like experiences | -0.235 | -0.304, -0.165 | 0.00407 | 5.22x10<sup>-11</sup> |
| Number of significantly distressing psychotic-like experiences | -0.762 | -0.989, -0.535 | 0.00408 | 4.78x10<sup>-11</sup> |
| **Intelligence PGS** | **Mean Difference** | **95% CI** | **R<sup>2</sup> of PGS** | **p** |
| Number of psychotic-like experiences | -0.280 | -0.392, -0.168 | 0.00229 | 8.98x10<sup>-7</sup> |
| Number of distressing psychotic-like experiences | -0.170 | -0.242, -0.0980 | 0.00202 | 3.83x10<sup>-6</sup> |
| Number of significantly distressing psychotic-like experiences | -0.589 | -0.823, -0.355 | 0.00230 | 8.01x10<sup>-7</sup> |

*Note:* R<sup>2</sup> of PGS refers to the partial variance explained by the respective PGS within the regression model
*Abbreviations:* ABCD = Adolescent Brain Cognitive Development; PGS = polygenic score; CI = confidence interval

Intelligence PGS was negatively associated with all psychotic-like experience outcomes in the regression models. A greater intelligence PGS was associated with decreased odds of psychotic-like experiences (OR: 0.829; 95% CI: 0.777–0.884; *p* = 1.35×10^-8^), distressing psychotic-like experiences (OR: 0.853; 95% CI: 0.800–0.909; *p* = 1.02×10^-6^), and significantly distressing psychotic-like experiences (OR: 0.795; 95% CI: 0.740–0.855; *p* = 6.32×10^-10^) (Figure 4), as well as the number of each type of psychotic-like experience reported (Table 4).

However, when the group classification analyses were carried out using the full SVM model, each performed at near the 0.5 level (AUROC: 0.550-0.613). Further detail available in the Supplementary Material. None of the group comparisons for either schizophrenia PGS or bipolar disorder PGS were significant after correction for multiple comparisons (*p*s > .00817).

## 4. Discussion

In this study, we aimed to examine the effect of polygenic scores for both schizophrenia and bipolar disorder on performance on specific cognitive domains, examine the effect of polygenic scores for intelligence and educational attainment on psychosis presentation, and extend previous research in this area by exploring these associations in both adults and children. Our results suggest that schizophrenia PGS and bipolar disorder PGS have different effects on cognitive performance, and that these effects differ across cognitive domains. We also found that intelligence PGS showed a stronger association with psychosis outcomes than educational attainment PGS in both adults and children.

### 4.1. Polygenic Scores for Psychosis on Cognitive Performance

Our evidence supports the previously reported negative association between schizophrenia PGS and cognitive performance (22, 38, 39). However, we showed this effect appeared to differ between adults and children, and between cognitive domains. In the adult sample, higher schizophrenia PGS was associated with poorer visuospatial processing performance, but not working memory or episodic memory. In the sample of children, a higher schizophrenia PGS was associated with poorer language ability, working memory, processing speed, and episodic memory, while there was no evidence of association with visuospatial processing. Methodological factors may have contributed to this difference (e.g. differences in the tests used, different sample sizes, controlling for clinical groups status in the PEIC analyses), but there is a possibility these findings represent a difference in the effect of schizophrenia risk over time or distinct maturational paths of the individual cognitive domains. However, alongside visuospatial processing, the block design task can be seen as a test of problem solving (e.g. (40)), which is in line with the negative effect of schizophrenia PGS on fluid intelligence in the ABCD Study^®^ sample. There is conflicting evidence in the literature on the effect of schizophrenia PGS on the other domains that differ between the two samples (28, 40, 73, 74). Further research is needed to pinpoint these associations. It is also worth noting that even the largest effect of the schizophrenia PGS on cognition was well below one standard deviation, the average deficits observed in up to 80% of people with psychosis (4). This suggests that while genetic factors may contribute to the deficit, the observed effect is modest and other factors must be involved.

In both samples, we found only weak evidence that polygenic scores for bipolar disorder negatively impact cognitive performance (27, 31–33, 75). There was only one test that showed a stronger association with bipolar disorder PGS than schizophrenia PGS: the Cash Choice task (though this did not pass the corrected significance threshold). Bipolar disorder PGS was associated with greater odds of choosing immediate gratification compared to delayed gratification, suggesting greater impulsivity. Impulsivity has been linked with bipolar disorder specifically (76, 77) so may serve as a mania/bipolar disorder-specific endophenotype, though other evidence suggests it is associated with psychosis more generally (78).

### 4.2. Polygenic Scores for Cognition on Psychosis Presentation

When looking at the effect of cognition-related polygenic scores on psychosis presentation, intelligence PGS showed a stronger association than educational attainment PGS. Intelligence PGS distinguished patients from controls in the adult sample (though with a much smaller effect size than that of schizophrenia or bipolar disorder PGS), and was negatively associated with all categories of psychotic-like experiences in the child sample. Educational attainment PGS showed a similar negative association with some categories of psychotic-like experiences, but did not distinguish between any clinical groups in the adult sample. These differences align with previous findings that polygenic scores for cognitive performance are more associated with schizophrenia case-status than educational attainment polygenic scores (35, 36, 49), possibly due to the non-genetic factors also involved in educational attainment (50, 79, 80).

One possible explanation for this relationship between cognition-related polygenic scores and psychosis presentation may be rooted in the degree of cognitive reserve that an individual has (81). It may be that individuals with lower cognition-related polygenic scores have a reduced cognitive reserve, which may in turn affect a range of factors should they develop symptoms of psychosis in adulthood. Cognition-related polygenic scores may therefore act as a moderator on the relationship between genetic risk for psychosis and symptom presentation/diagnosis in adulthood.

Although the Prodromal Questionnaire–Brief Child Version (PQ-BC) has been validated as a measure of early risk for psychosis (82), these symptoms are not necessarily indicative of psychosis in adulthood (83). In this sample, the proportion of participants that reported experiencing at least one psychotic-like experience was well over the proportion expected to develop psychosis (Table 3) (and it should also be noted that a formal, current diagnosis of schizophrenia was an exclusion criteria in the ABCD Study^®^). This may explain the reduced effect of cognition-related PGSs when distinguishing between adults meeting the diagnostic criteria for a psychotic disorder. Psychotic-like experiences may instead be indicative of an increased risk of later psychopathology more generally (84). In terms of psychosis risk specifically, it may be better to focus on those who experience multiple distressing psychotic-like experiences, or those who report such experiences at multiple timepoints (e.g. (85)).

### 4.3. Limitations

The method used to calculate the polygenic scores, PRS-CSx, provides improved accuracy in polygenic prediction compared to previous methods, by allowing summary statistics from multiple ancestries to be used together (86). However, the accuracy is still below what would be needed for clinical practice and the need for an ancestry reference panel limits the application of polygenic scores in admixed/underrepresented populations (59, 87).

While we made efforts to improve the ancestry diversity of the samples we included, the accuracy of the polygenic score calculations are still limited by the GWAS discovery samples, which remain mostly European (61–64). Efforts are in place to carry out GWASs in global populations beyond Europe (88). The Human Pangenome Reference Consortium (89) is working to create a reference panel that better represents diverse populations; and the Polygenic RIsk MEthods in Diverse populations (PRIMED) Consortium (90) seeks to improve applicability of PGSs to diverse populations.

Another limitation is that PGSs only account for common variants associated with an outcome. This is seen by the SNP heritability for schizophrenia, bipolar disorder, and intelligence, each reported as around 20% (61, 62, 64), despite overall heritability rates of 60-80% for schizophrenia (61), 60-85% for bipolar disorder (62), and 50% for intelligence (64). Other genetic factors, including copy number variants (CNVs), are known to be associated with psychosis and cognitive functioning (91, 92). However, there is evidence that, at the population level, common variants may play a larger role in health- and cognition-related outcomes than CNVs (93).

### 4.4. Implications

As cognitive impairment in psychosis is associated with poorer health outcomes (4, 94, 95), evidence that this association is seen at the neurobiological level may help to better identify those at risk. At present, polygenic scores are not accurate enough for clinical use (96). However, they remain a useful research tool. Despite limitations, introducing genomics into psychiatric care may provide another avenue for identifying those who would benefit from early intervention. The combination of high genetic risk for psychosis and polygenic scores for lower cognitive functioning may be useful for stratification and personalised treatments.

These results, if replicated, add to the growing evidence for a genetic component in the relationship between psychosis and cognitive impairment (22). The focus should be on replicating this research in different ancestry groups. As polygenic profiling becomes more widely available, without these efforts, such techniques may further exacerbate health disparities/inequity instead of improving healthcare (97, 98).

Using individual cognitive domains as the exposure, rather than broader measures of cognitive performance, may help to further pinpoint the cognitive domains most associated with psychosis. There is currently a lack of research into such genetic associations; the largest GWAS currently available is for reaction time, which found 2,022 associated variants over 42 loci (99). There are also a number of other ways to use the psychosis-related exposures, for example: looking at only variants associated with a specific disorder (e.g. those associated with schizophrenia only, as opposed to those that also overlap with bipolar disorder; (29, 31)).

Finally, longitudinal studies such as the ABCD Study^®^ provide the opportunity for developmental research in this field (100). This allows patterns to be followed across the developmental period and to discover possible associations with sustained psychotic-like experiences (85), diagnosis, or cognitive deficits. This also covers the period previously used to examine the change in heritability of cognitive functioning (101), so this sample could be used to examine whether this change affects the impact of genetic factors on the relationship between psychosis and cognition.

### 4.5. Conclusion

We found evidence that genetic variants associated with both psychosis (specifically, schizophrenia) and cognition (educational attainment and intelligence) are implicated in the relationship between psychosis and cognitive impairment. This supports the use of cognitive function as an endophenotype for psychosis. The different effects of schizophrenia risk on performance within individual cognitive domains suggest that specific domains may serve as better endophenotypes than overall cognitive functioning.

## Data Availability

All data produced in the present study are available upon reasonable request to the authors

## Acknowledgements

Collaborators of Psychosis Endophenotypes International Consortium (PEIC) include: Andrew M McIntosh; Cathryn Lewis; John Powell; Dan Rujescu; Matthias Weisbrod; Ina Giegling; Maria J. Arranz; Marta Di Forti; and Kuang Lin. The Santander cohort was supported by Instituto de Salud Carlos III (PI020499, PI050427, PI060507), SENY Fundació (CI 2005-0308007), Fundacion Ramón Areces and Fundacion Marqués de Valdecilla (API07/011, API10/13). Grant ISCIII: ICI21_00089 and PI22_01379.

Genetic Risk and Outcome of Psychosis (GROUP) Investigators includes the following people: Behrooz Z. Alizadeh^1,2^ Therese van Amelsvoort^3^ Lieuwe de Haan^4,5^ Marieke van der Pluijm^4,5^ Claudia J.P. Simons^3,6^ Wim Veling^2^

^1^Department of Epidemiology, University Medical Center Groningen, University of Groningen, Groningen, The Netherlands

^2^Department of Psychiatry, University Medical Center Groningen, University of Groningen, Groningen, The Netherlands

^3^Department of Psychiatry and Neuropsychology, Maastricht University Medical Center, School for Mental Health and Neuroscience, Maastricht, The Netherlands

^4^Department of Psychiatry, Amsterdam UMC, University of Amsterdam, The Netherlands

^5^Arkin Institute for Mental Health, Amsterdam, The Netherlands

^6^ GGzE Institute for Mental Health Care, Eindhoven, The Netherlands Data used in the preparation of this article were obtained from the Adolescent Brain Cognitive Development^SM^ (ABCD) Study (https://abcdstudy.org), held in the NIMH Data Archive (NDA). This is a multisite, longitudinal study designed to recruit more than 10,000 children age 9-10 and follow them over 10 years into early adulthood. The ABCD Study^®^ is supported by the National Institutes of Health and additional federal partners under award numbers U01DA041048, U01DA050989, U01DA051016, U01DA041022, U01DA051018, U01DA051037, U01DA050987, U01DA041174, U01DA041106, U01DA041117, U01DA041028, U01DA041134, U01DA050988, U01DA051039, U01DA041156, U01DA041025, U01DA041120, U01DA051038, U01DA041148, U01DA041093, U01DA041089, U24DA041123, U24DA041147. A full list of supporters is available at https://abcdstudy.org/federal-partners.html. A listing of participating sites and a complete listing of the study investigators can be found at https://abcdstudy.org/consortium_members/. ABCD consortium investigators designed and implemented the study and/or provided data but did not necessarily participate in the analysis or writing of this report. This manuscript reflects the views of the authors and may not reflect the opinions or views of the NIH or ABCD consortium investigators.

The ABCD data repository grows and changes over time. The ABCD data used in this report came from doi:10.15154/kwr5-qf75. DOIs can be found at https://dx.doi.org/10.15154/kwr5-qf75. L.V. is supported by Medical Research Council (MR/W020238/1). E.B. is supported by Medical Research Council (G1100583, MR/W020238/1); National Institute of Health Research (NIHR200756); Mental Health Research UK - John Grace QC Scholarship 2018; Economic Social Research Council’s Co-funded doctoral award; The British Medical Association’s Margaret Temple Fellowship; Medical Research Council New Investigator and Centenary Awards (G0901310, G1100583); NIHR Biomedical Research Centre at University College London Hospitals NHS Foundation Trust and University College London (NIHR BRC UCL). K.J.’s work was undertaken as part of the UCL-Birkbeck MRC-DTP PhD program, generously funded by the Medical Research Council (MR/W006774/1). A.R.B. is funded by the Wellcome Trust through a PhD Fellowship in Mental Health Science (218497/Z/19/Z). This research was funded in whole or in part by the Wellcome Trust. For the purpose of Open Access, the author has applied a CC BY public copyright licence to any Author Accepted Manuscript (AAM) version arising from this submission. N.S.K. is supported by a studentship co-funded by the Economic and Social Research Council (ESRC) and Mental Health Research UK (ES/P000592/1). D.P. received the FCT 2022.00586.CEECIND felllowship (DOI: 10.54499/2022.00586.CEECIND/CP1722/CT0011). All other authors report no biomedical financial interests or potential conflicts of interest.

## Disclosures

S.B. has received financial support for symposia from Medice, Takeda and is on the advisory board for AGB-Pharma. All other authors report no disclosures.

## Notes

### Author Declarations

Ethical approval granted by the National Health Service Multicentre Research Ethics Committee (references 03/11/090 and 19/LO/1403). Identifiable data are stored in a secure system at University College London that meets the requirements of NHS Information Governance Toolkit, ISO 27001, and all national policies.

## References

1. Anderson IM, Haddad PM, Scott J. Bipolar disorder. BMJ. 2012;345:e8508.

2. Hilker R, Helenius D, Fagerlund B, Skytthe A, Christensen K, Werge TM, et al. Heritability of Schizophrenia and Schizophrenia Spectrum Based on the Nationwide Danish Twin Register. Biol Psychiatry. 2018;83(6):492–8.

3. National Health Service (NHS). Overview - Psychosis 2019 [Available from: https://www.nhs.uk/mental-health/conditions/psychosis/overview/.

4. McCleery A, Nuechterlein KH. Cognitive impairment in psychotic illness: prevalence, profile of impairment, developmental course, and treatment considerations□Dialogues Clin Neurosci. 2019;21(3):239–48.

5. Vohringer PA, Barroilhet SA, Amerio A, Reale ML, Alvear K, Vergne D, Ghaemi SN. Cognitive impairment in bipolar disorder and schizophrenia: a systematic review. Front Psychiatry. 2013;4:87.

6. East-Richard C, R. -Mercier A, Nadeau D, Cellard C. Transdiagnostic neurocognitive deficits in psychiatry: A review of meta-analyses. Canadian Psychology / Psychologie canadienne. 2020;61(3):190–214.

7. Schwarz E, Tost H, Meyer-Lindenberg A. Working memory genetics in schizophrenia and related disorders: An RDoC perspective. Am J Med Genet B Neuropsychiatr Genet. 2016;171B(1):121–31.

8. Ma X, Wang Q, Sham PC, Liu X, Rabe-Hesketh S, Sun X, et al. Neurocognitive deficits in first-episode schizophrenic patients and their first-degree relatives. Am J Med Genet B Neuropsychiatr Genet. 2007;144B(4):407–16.

9. Hoff AL, Svetina C, Maurizio AM, Crow TJ, Spokes K, DeLisi LE. Familial cognitive deficits in schizophrenia. Am J Med Genet B Neuropsychiatr Genet. 2005;133B(1):43–9.

10. Snitz BE, Macdonald AW, 3rd, Carter CS. Cognitive deficits in unaffected first-degree relatives of schizophrenia patients: a meta-analytic review of putative endophenotypes. Schizophr Bull. 2006;32(1):179–94.

11. Blakey R, Ranlund S, Zartaloudi E, Cahn W, Calafato S, Colizzi M, et al. Associations between psychosis endophenotypes across brain functional, structural, and cognitive domains. Psychol Med. 2018;48(8):1325–40.

12. Catalan A, Salazar de Pablo G, Aymerich C, Damiani S, Sordi V, Radua J, et al. Neurocognitive Functioning in Individuals at Clinical High Risk for Psychosis: A Systematic Review and Meta-analysis. JAMA Psychiatry. 2021;78(8):859–67.

13. Seidman LJ, Giuliano AJ, Smith CW, Stone WS, Glatt SJ, Meyer E, et al. Neuropsychological functioning in adolescents and young adults at genetic risk for schizophrenia and affective psychoses: results from the Harvard and Hillside Adolescent High Risk Studies. Schizophr Bull. 2006;32(3):507–24.

14. Plomin R, von Stumm S. The new genetics of intelligence. Nat Rev Genet. 2018;19(3):148–59.

15. Blokland GAM, Mesholam-Gately RI, Toulopoulou T, Del Re EC, Lam M, DeLisi LE, et al. Heritability of Neuropsychological Measures in Schizophrenia and Nonpsychiatric Populations: A Systematic Review and Meta-analysis. Schizophr Bull. 2017;43(4):788–800.

16. Antila M, Tuulio-Henriksson A, Kieseppa T, Soronen P, Palo OM, Paunio T, et al. Heritability of cognitive functions in families with bipolar disorder. Am J Med Genet B Neuropsychiatr Genet. 2007;144B(6):802–8.

17. Gottesman II, Gould TD. The Endophenotype Concept in Psychiatry: Etymology and Strategic Intentions. American Journal of Psychiatry. 2003;160(4):636–45.

18. Gould TD, Gottesman, II. Psychiatric endophenotypes and the development of valid animal models. Genes Brain Behav. 2006;5(2):113–9.

19. Gur RE, Calkins ME, Gur RC, Horan WP, Nuechterlein KH, Seidman LJ, Stone WS. The Consortium on the Genetics of Schizophrenia: neurocognitive endophenotypes. Schizophr Bull. 2007;33(1):49–68.

20. Toulopoulou T, Zhang X, Cherny S, Dickinson D, Berman KF, Straub RE, et al. Polygenic risk score increases schizophrenia liability through cognition-relevant pathways. Brain. 2019;142(2):471–85.

21. Fullerton JM, Nurnberger JI. Polygenic risk scores in psychiatry: Will they be useful for clinicians? F1000Res. 2019;8.

22. Mallet J, Le Strat Y, Dubertret C, Gorwood P. Polygenic Risk Scores Shed Light on the Relationship between Schizophrenia and Cognitive Functioning: Review and Meta-Analysis. J Clin Med. 2020;9(2).

23. Richards AL, Pardinas AF, Frizzati A, Tansey KE, Lynham AJ, Holmans P, et al. The Relationship Between Polygenic Risk Scores and Cognition in Schizophrenia. Schizophr Bull. 2020;46(2):336–44.

24. Schaupp SK, Schulze TG, Budde M. Let’s Talk about the Association between Schizophrenia Polygenic Risk Scores and Cognition in Patients and the General Population: A Review. Journal of Psychiatry and Brain Science. 2018;3(6):12.

25. Habtewold TD, Liemburg EJ, Islam MA, de Zwarte SMC, Boezen HM, Investigators G, et al. Association of schizophrenia polygenic risk score with data-driven cognitive subtypes: A six-year longitudinal study in patients, siblings and controls. Schizophr Res. 2020;223:135–47.

26. Mistry S, Harrison JR, Smith DJ, Escott-Price V, Zammit S. The use of polygenic risk scores to identify phenotypes associated with genetic risk of schizophrenia: Systematic review. Schizophr Res. 2018;197:2–8.

27. Comes AL, Senner F, Budde M, Adorjan K, Anderson-Schmidt H, Andlauer TFM, et al. The genetic relationship between educational attainment and cognitive performance in major psychiatric disorders. Transl Psychiatry. 2019;9(1):210.

28. Engen MJ, Lyngstad SH, Ueland T, Simonsen CE, Vaskinn A, Smeland O, et al. Polygenic scores for schizophrenia and general cognitive ability: associations with six cognitive domains, premorbid intelligence, and cognitive composite score in individuals with a psychotic disorder and in healthy controls. Transl Psychiatry. 2020;10(1):416.

29. Ohi K, Nishizawa D, Sugiyama S, Takai K, Kuramitsu A, Hasegawa J, et al. Polygenic Risk Scores Differentiating Schizophrenia From Bipolar Disorder Are Associated With Premorbid Intelligence in Schizophrenia Patients and Healthy Subjects. Int J Neuropsychopharmacol. 2021;24(7):562–9.

30. Escott-Price V, Bracher-Smith M, Menzies G, Walters J, Kirov G, Owen MJ, O’Donovan MC. Genetic liability to schizophrenia is negatively associated with educational attainment in UK Biobank. Mol Psychiatry. 2020;25(4):703–5.

31. Mistry S, Escott-Price V, Florio AD, Smith DJ, Zammit S. Investigating associations between genetic risk for bipolar disorder and cognitive functioning in childhood. J Affect Disord. 2019;259:112–20.

32. Liebers DT, Pirooznia M, Seiffudin F, Musliner KL, Zandi PP, Goes FS. Polygenic Risk of Schizophrenia and Cognition in a Population-Based Survey of Older Adults. Schizophr Bull. 2016;42(4):984–91.

33. Ranlund S, Calafato S, Thygesen JH, Lin K, Cahn W, Crespo-Facorro B, et al. A polygenic risk score analysis of psychosis endophenotypes across brain functional, structural, and cognitive domains. Am J Med Genet B Neuropsychiatr Genet. 2018;177(1):21–34.

34. Power RA, Steinberg S, Bjornsdottir G, Rietveld CA, Abdellaoui A, Nivard MM, et al. Polygenic risk scores for schizophrenia and bipolar disorder predict creativity. Nat Neurosci. 2015;18(7):953–5.

35. Shafee R, Nanda P, Padmanabhan JL, Tandon N, Alliey-Rodriguez N, Kalapurakkel S, et al. Polygenic risk for schizophrenia and measured domains of cognition in individuals with psychosis and controls. Transl Psychiatry. 2018;8(1):78.

36. Sorensen HJ, Debost JC, Agerbo E, Benros ME, McGrath JJ, Mortensen PB, et al. Polygenic Risk Scores, School Achievement, and Risk for Schizophrenia: A Danish Population-Based Study. Biol Psychiatry. 2018;84(9):684–91.

37. Legge SE, Cardno AG, Allardyce J, Dennison C, Hubbard L, Pardinas AF, et al. Associations Between Schizophrenia Polygenic Liability, Symptom Dimensions, and Cognitive Ability in Schizophrenia. JAMA Psychiatry. 2021;78(10):1143–51.

38. He Q, Jantac Mam-Lam-Fook C, Chaignaud J, Danset-Alexandre C, Iftimovici A, Gradels Hauguel J, et al. Influence of polygenic risk scores for schizophrenia and resilience on the cognition of individuals at-risk for psychosis. Transl Psychiatry. 2021;11(1):518.

39. Ohi K, Takai K, Kuramitsu A, Sugiyama S, Soda M, Kitaichi K, Shioiri T. Causal associations of intelligence with schizophrenia and bipolar disorder: A Mendelian randomization analysis. Eur Psychiatry. 2021;64(1):e61.

40. Hubbard L, Tansey KE, Rai D, Jones P, Ripke S, Chambert KD, et al. Evidence of Common Genetic Overlap Between Schizophrenia and Cognition. Schizophr Bull. 2016;42(3):832–42.

41. Lencz T, Knowles E, Davies G, Guha S, Liewald DC, Starr JM, et al. Molecular genetic evidence for overlap between general cognitive ability and risk for schizophrenia: a report from the Cognitive Genomics consorTium (COGENT). Mol Psychiatry. 2014;19(2):168–74.

42. McGrath JJ, Féron FP, Burne THJ, Mackay-Sim A, Eyles DW. The neurodevelopmental hypothesis of schizophrenia: a review of recent developments. Annals of Medicine. 2003;35(2):86–93.

43. Murray RM, Bhavsar V, Tripoli G, Howes O. 30 Years on: How the Neurodevelopmental Hypothesis of Schizophrenia Morphed Into the Developmental Risk Factor Model of Psychosis. Schizophrenia Bulletin. 2017;43(6):1190–6.

44. Fusar-Poli P, Deste G, Smieskova R, Barlati S, Yung AR, Howes O, et al. Cognitive functioning in prodromal psychosis: a meta-analysis. Arch Gen Psychiatry. 2012;69(6):562–71.

45. Liu Y, Wang G, Jin H, Lyu H, Liu Y, Guo W, et al. Cognitive deficits in subjects at risk for psychosis, first-episode and chronic schizophrenia patients. Psychiatry Res. 2019;274:235–42.

46. Tor J, Dolz M, Sintes-Estevez A, de la Serna E, Puig O, Munoz-Samons D, et al. Neuropsychological profile of children and adolescents with psychosis risk syndrome: the CAPRIS study. Eur Child Adolesc Psychiatry. 2020;29(9):1311–24.

47. Bora E, Murray RM. Meta-analysis of cognitive deficits in ultra-high risk to psychosis and first-episode psychosis: do the cognitive deficits progress over, or after, the onset of psychosis? Schizophr Bull. 2014;40(4):744–55.

48. Karcher NR, Loewy RL, Savill M, Avenevoli S, Huber RS, Simon TJ, et al. Replication of Associations With Psychotic-Like Experiences in Middle Childhood From the Adolescent Brain Cognitive Development (ABCD) Study. Schizophr Bull Open. 2020;1(1):sgaa009.

49. Karcher NR, Paul SE, Johnson EC, Hatoum AS, Baranger DAA, Agrawal A, et al. Psychotic-like Experiences and Polygenic Liability in the Adolescent Brain Cognitive Development Study. Biol Psychiatry Cogn Neurosci Neuroimaging. 2022;7(1):45–55.

50. Park J, Lee E, Cho G, Hwang H, Joo YY, Cha J. Gene-Environment Causal Pathway to Impoverished Cognitive Development Contributes to Psychotic-Like Experience in Children. medRxiv. 2021:2021.12.28.21268440.

51. Bramon E, Pirinen M, Strange A, Lin K, Freeman C, Bellenguez C, et al. A genome-wide association analysis of a broad psychosis phenotype identifies three loci for further investigation. Biol Psychiatry. 2014;75(5):386–97.

52. ABCD Study. About the Study 2021 [Available from: https://abcdstudy.org/about/.

53. Wang B, Giannakopoulou O, Austin-Zimmerman I, Irizar H, Harju-Seppanen J, Zartaloudi E, et al. Adolescent Verbal Memory as a Psychosis Endophenotype: A Genome-Wide Association Study in an Ancestrally Diverse Sample. Genes (Basel). 2022;13(1).

54. Uban KA, Horton MK, Jacobus J, Heyser C, Thompson WK, Tapert SF, et al. Biospecimens and the ABCD study: Rationale, methods of collection, measurement and early data. Dev Cogn Neurosci. 2018;32:97–106.

55. Gogarten SM, Sofer T, Chen H, Yu C, Brody JA, Thornton TA, et al. Genetic association testing using the GENESIS R/Bioconductor package. Bioinformatics. 2019;35(24):5346–8.

56. R Core Team. R: A language and environment for statistical computing. R Foundation for Statistical Computing, Vienna, Austria. 2020 [Available from: https://www.R-project.org/.

57. Manichaikul A, Mychaleckyj JC, Rich SS, Daly K, Sale M, Chen WM. Robust relationship inference in genome-wide association studies. Bioinformatics. 2010;26(22):2867–73.

58. Zheng X, Levine D, Shen J, Gogarten SM, Laurie C, Weir BS. A high-performance computing toolset for relatedness and principal component analysis of SNP data. Bioinformatics. 2012;28(24):3326–8.

59. Ruan Y, Lin Y-F, Feng Y-CA, Chen C-Y, Lam M, Guo Z, et al. Improving polygenic prediction in ancestrally diverse populations. Nature Genetics. 2022;54(5):573–80.

60. Auton A, Abecasis GR, Altshuler DM, Durbin RM, Abecasis GR, Bentley DR, et al. A global reference for human genetic variation. Nature. 2015;526(7571):68–74.

61. Trubetskoy V, Pardinas AF, Qi T, Panagiotaropoulou G, Awasthi S, Bigdeli TB, et al. Mapping genomic loci implicates genes and synaptic biology in schizophrenia. Nature. 2022;604(7906):502–8.

62. Mullins N, Forstner AJ, O’Connell KS, Coombes B, Coleman JRI, Qiao Z, et al. Genome-wide association study of more than 40,000 bipolar disorder cases provides new insights into the underlying biology. Nat Genet. 2021;53(6):817–29.

63. Lee JJ, Wedow R, Okbay A, Kong E, Maghzian O, Zacher M, et al. Gene discovery and polygenic prediction from a genome-wide association study of educational attainment in 1.1 million individuals. Nat Genet. 2018;50(8):1112–21.

64. Savage JE, Jansen PR, Stringer S, Watanabe K, Bryois J, de Leeuw CA, et al. Genome-wide association meta-analysis in 269,867 individuals identifies new genetic and functional links to intelligence. Nat Genet. 2018;50(7):912–9.

65. Wechsler D. WAIS-R: Wechsler adult intelligence scale-revised. New York, N.Y.: Psychological Corporation, [1981] ©1981; 1981.

66. Wechsler D. Wechsler Adult Intelligence Scale (3rd ed.). 1997.

67. Rey A. L’examen clinique en psychologie. [The clinical examination in psychology.]. Oxford, England: Presses Universitaries De France; 1958. 222-p.

68. Luciana M, Bjork JM, Nagel BJ, Barch DM, Gonzalez R, Nixon SJ, Banich MT. Adolescent neurocognitive development and impacts of substance use: Overview of the adolescent brain cognitive development (ABCD) baseline neurocognition battery. Dev Cogn Neurosci. 2018;32:67–79.

69. Soto T, Kraper C. Block Design Subtest. In: Volkmar FR, editor. Encyclopedia of Autism Spectrum Disorders. New York, NY: Springer New York; 2013. p. 464–5.

70. Wambach D, Lamar M, Swenson R, Penney DL, Kaplan E, Libon DJ. Digit Span. In: Kreutzer JS, DeLuca J, Caplan B, editors. Encyclopedia of Clinical Neuropsychology. New York, NY: Springer New York; 2011. p. 844–9.

71. Bracher-Smith M, Crawford K, Escott-Price V. Machine learning for genetic prediction of psychiatric disorders: a systematic review. Mol Psychiatry. 2021;26(1):70–9.

72. Breiman L. Random Forests. Machine Learning. 2001;45(1):5–32.

73. Nakahara S, Medland S, Turner JA, Calhoun VD, Lim KO, Mueller BA, et al. Polygenic risk score, genome-wide association, and gene set analyses of cognitive domain deficits in schizophrenia. Schizophr Res. 2018;201:393–9.

74. Koch E, Nyberg L, Lundquist A, Pudas S, Adolfsson R, Kauppi K. Sex-specific effects of polygenic risk for schizophrenia on lifespan cognitive functioning in healthy individuals. Transl Psychiatry. 2021;11(1):520.

75. Mistry S, Harrison JR, Smith DJ, Escott-Price V, Zammit S. The use of polygenic risk scores to identify phenotypes associated with genetic risk of bipolar disorder and depression: A systematic review. J Affect Disord. 2018;234:148–55.

76. Strakowski SM, Fleck DE, DelBello MP, Adler CM, Shear PK, Kotwal R, Arndt S. Impulsivity across the course of bipolar disorder. Bipolar Disord. 2010;12(3):285–97.

77. Reddy LF, Lee J, Davis MC, Altshuler L, Glahn DC, Miklowitz DJ, Green MF. Impulsivity and risk taking in bipolar disorder and schizophrenia. Neuropsychopharmacology. 2014;39(2):456–63.

78. Ahn WY, Rass O, Fridberg DJ, Bishara AJ, Forsyth JK, Breier A, et al. Temporal discounting of rewards in patients with bipolar disorder and schizophrenia. J Abnorm Psychol. 2011;120(4):911–21.

79. Smith-Woolley E, Selzam S, Plomin R. Polygenic score for educational attainment captures DNA variants shared between personality traits and educational achievement. Journal of Personality and Social Psychology. 2019;117(6):1145–63.

80. Ujma PP, Eszlari N, Millinghoffer A, Bruncsics B, Torok D, Petschner P, et al. Genetic effects on educational attainment in Hungary. Brain Behav. 2022;12(1):e2430.

81. Ayesa-Arriola R, de la Foz VO, Murillo-Garcia N, Vazquez-Bourgon J, Juncal-Ruiz M, Gomez-Revuelta M, et al. Cognitive reserve as a moderator of outcomes in five clusters of first episode psychosis patients: a 10-year follow-up study of the PAFIP cohort. Psychol Med. 2023;53(5):1891–905.

82. Karcher NR, Barch DM, Avenevoli S, Savill M, Huber RS, Simon TJ, et al. Assessment of the Prodromal Questionnaire-Brief Child Version for Measurement of Self-reported Psychoticlike Experiences in Childhood. JAMA Psychiatry. 2018;75(8):853–61.

83. Karcher NR, Perino MT, Barch DM. An item response theory analysis of the Prodromal Questionnaire-Brief Child Version: Developing a screening form that informs understanding of self-reported psychotic-like experiences in childhood. Journal of Abnormal Psychology. 2020;129(3):293–304.

84. Schimmelmann BG, Walger P, Schultze-Lutter F. The significance of at-risk symptoms for psychosis in children and adolescents. Can J Psychiatry. 2013;58(1):32–40.

85. Karcher NR, Loewy RL, Savill M, Avenevoli S, Huber RS, Makowski C, et al. Persistent and distressing psychotic-like experiences using adolescent brain cognitive developmentl7l study data. Mol Psychiatry. 2022;27(3):1490–501.

86. Ge T, Irvin MR, Patki A, Srinivasasainagendra V, Lin Y-F, Tiwari HK, et al. Development and validation of a trans-ancestry polygenic risk score for type 2 diabetes in diverse populations. Genome Medicine. 2022;14(1):70.

87. Ge T, Chen CY, Ni Y, Feng YA, Smoller JW. Polygenic prediction via Bayesian regression and continuous shrinkage priors. Nat Commun. 2019;10(1):1776.

88. Duncan L, Shen H, Gelaye B, Meijsen J, Ressler K, Feldman M, et al. Analysis of polygenic risk score usage and performance in diverse human populations. Nature Communications. 2019;10(1):3328.

89. Human Pangenome Reference Consortium (HPRC). Human Pangenome Reference Consortium 2022 [Available from: https://humanpangenome.org/index.html.

90. National Human Genome Research Institute. Polygenic RIsk MEthods in Diverse populations (PRIMED) Consortium 2022 [Available from: https://www.genome.gov/Funded-Programs-Projects/PRIMED-Consortium.

91. Thygesen JH, Presman A, Harju-Seppanen J, Irizar H, Jones R, Kuchenbaecker K, et al. Genetic copy number variants, cognition and psychosis: a meta-analysis and a family study. Mol Psychiatry. 2021;26(9):5307–19.

92. Hubbard L, Rees E, Morris DW, Lynham AJ, Richards AL, Pardinas AF, et al. Rare Copy Number Variants Are Associated With Poorer Cognition in Schizophrenia. Biol Psychiatry. 2021;90(1):28–34.

93. Saarentaus EC, Havulinna AS, Mars N, Ahola-Olli A, Kiiskinen TTJ, Partanen J, et al. Polygenic burden has broader impact on health, cognition, and socioeconomic outcomes than most rare and high-risk copy number variants. Mol Psychiatry. 2021;26(9):4884–95.

94. Allott K, Liu P, Proffitt TM, Killackey E. Cognition at illness onset as a predictor of later functional outcome in early psychosis: systematic review and methodological critique. Schizophr Res. 2011;125(2-3):221–35.

95. Song J, Yao S, Kowalec K, Lu Y, Sariaslan A, Szatkiewicz JP, et al. The impact of educational attainment, intelligence and intellectual disability on schizophrenia: a Swedish population-based register and genetic study. Mol Psychiatry. 2022;27(5):2439–47.

96. Fries GR. Polygenic risk scores and their potential clinical use in psychiatry: are we there yet? Braz J Psychiatry. 2020;42(5):459–60.

97. Martin AR, Kanai M, Kamatani Y, Okada Y, Neale BM, Daly MJ. Clinical use of current polygenic risk scores may exacerbate health disparities. Nat Genet. 2019;51(4):584–91.

98. Palk AC, Dalvie S, de Vries J, Martin AR, Stein DJ. Potential use of clinical polygenic risk scores in psychiatry - ethical implications and communicating high polygenic risk. Philos Ethics Humanit Med. 2019;14(1):4.

99. Davies G, Lam M, Harris SE, Trampush JW, Luciano M, Hill WD, et al. Study of 300,486 individuals identifies 148 independent genetic loci influencing general cognitive function. Nat Commun. 2018;9(1):2098.

100. Barch DM, Albaugh MD, Avenevoli S, Chang L, Clark DB, Glantz MD, et al. Demographic, physical and mental health assessments in the adolescent brain and cognitive development study: Rationale and description. Dev Cogn Neurosci. 2018;32:55–66.

101. Haworth CM, Wright MJ, Luciano M, Martin NG, de Geus EJ, van Beijsterveldt CE, et al. The heritability of general cognitive ability increases linearly from childhood to young adulthood. Mol Psychiatry. 2010;15(11):1112–20.

