## Supplementary Material for "Effect of Polygenic Scores on the Relationship Between Psychosis and Cognitive Performance"

**Genotyping, Quality Control, and Imputation**

***Psychosis Endophenotypes International Consortium (PEIC)***

A total of 6,935 blood samples, collected from across the consortium sites, were sent to the Wellcome Trust Sanger Institute (Cambridge, UK). Samples were processed in a 96-well plate format, with each plate carrying a positive and negative control. DNA concentrations were quantified using a PicoGreen assay (Invitrogen, Life Technologies, Grand Island, New York) and an aliquot assayed by agarose gel electrophoresis. To pass quality control, DNA concentrations could not be degraded and had to be at least 50 ng/mL. Thirty SNPs, including sex chromosome markers, were typed on the Sequenom platform and then entered whole genome genotyping. 347 samples were excluded due to degradation, insufficient DNA, or incorrect sex classification. Remaining samples were then sent to the Affymetrix Services Laboratory ([www.affymetrix.com](http://www.affymetrix.com)).

Samples were genotyped using the Genome-wide Human SNP Array. Genotypes were called using the CHIAMO algorithm that was modified for use with the Affymetrix 6.0 genotyping array (1, 2). Around 30% of the samples were excluded due to poor signal-to-noise ratio in the genotyping array; those excluded were equally distributed across the clinical groups (32% of patients, 30% of relatives and 30% of controls; χ2 (2 df) = 3.2; *p* = 0.20).

Standard quality control was carried out at UCL using PEDSTATS (3), Evoker (4), LDAK (5), and PLINK (6). In total, 234,363 SNPS were excluded (7):

- 11,610 due to a study-wide missing rate of > 5%
- 26,858 due to four or more Mendelian inheritance errors
- 2,404 due to departure from Hardy-Weinberg equilibrium (*p* < 10^–6^)
- 145,097 due to a MAF < 0.02
- 38,895 from the sex chromosomes or mitochondrial DNA
- 9,499 after visual inspection of genotyping intensity plots

Quality controlled genotypes were then uploaded to the Sanger Imputation Sever (<https://imputation.sanger.ac.uk>). Pre-phasing and imputation were conducted according to the EAGLE2/PWBT pipeline, based on the Haplotype Reference Consortium panel (r1.1) (8, 9). Imputed genotypes were then converted into best-guess format using a heard-cell threshold of 0.8; SNPs with an INFO score < 0.8 were excluded. Additional quality control was then conducted on these imputed genotypes using PLINK (6) to exclude SNPs with a missing rate of > 5%, a MAF < 1%, departure from the Hardy-Weinberg equilibrium (*p* < 1x10^-6^), Mendelian error rate > 10%, or cases versus controls data missingness significance < 5x10^-6^. Samples were excluded due to a missing rate > 5%, a Mendelian error rate > 5%, or an inbreeding coefficient > 0.1. LDAK (5) was used to identify duplicates or twins; one of each pair was removed.

In total, 6,215,801 SNPs and 4,835 samples remained after quality control.

***Adolescent Brain Cognitive Development (ABCD) Study***

DNA was extracted from blood or saliva samples that were shipped to the Rutgers University Cell and DNA Repository (RUCDR; New Jersey, USA). The Affymetrix NIDA Smokescreen Array was used to genotype samples (10), which contained 733,293 SNPs. DNA was extracted using the Chemagen based/Chemagic STAR DNA Saliva4k Kit (CMG-1755-A). Genotyping was done using the Affimetrix GeneTital Instrument. Samples from either blood or saliva samples were used, whichever had successful call rates, a higher non-missing rate, matched genetic sex, or less extensive identity by state (11).

RUCDR performed quality control based on calling signals and variant call rates. Study-based quality control was then conducted by the ABCD Data Analysis, Informatics & Resource Center (DAIRC), following the RICOPILI pipeline (12). Samples were excluded with a call rate < .09 or sex violations; SNPs were removed with a call rate of < 0.9. This resulted in a sample of 11,099 participants and 516,598 genetic variants.

Imputation was conducted by the ABCD Study^®^ using the TOPMed imputation server. Pre-imputation steps were followed according to TOPMed recomemndations (<https://topmedimpute.readthedocs.io/en/latest/prepare-your-data.html>). Data were checked against reference panels (13, 14) then uploaded to the TOPMed Imputation Server and imputation was performed using mixed ancestry and Eagle v2.4 phasing (15, 16).

We downloaded this data from the ABCD Data Repository and used bcftools to annotate the rs IDs, based on dbSNP153 (17). Imputed SNPs with dosage levels were converted to best-guess genotype format using PLINK v2.0 with a hard call threshold of 0.1 (6).

Further, post-imputation quality control was then conducted to remove samples with one problematic plate (associated with failures advertised by the ABCD Study^®^), or SNPs with imputation quality scores (r2) < 0.3 or MAF < 0.01 (11). This left a sample of 11,017 participants 11,229,083 variants.

**Relationship Inference and Principal Component Analysis**

***Psychosis Endophenotypes International Consortium (PEIC) and Adolescent Brain Cognitive Development (ABCD) Study***

The GENESIS R/Bioconductor package (18, 19) was used to account for familial relatedness and population structure, by generating a kinship matrix and conducting a principal component (PC) analysis. Using the genotyped data that passed quality control, an unadjusted kinship matrix was generated using KING-robust 2.2.5 (20). The kinship coefficient is defined as the probability that two randomly selected alleles from two participants are identical by descent; the kinship coefficient for monozygotic twins is expected to be 0.5 (2^−2/2^) and that of first-degree relatives is expected to be 0.25 (2^−4/2^) (20). Using this, 0.354 (2^−3/2^) was used as the threshold for differentiating monozygotic twins and first-degree relatives; the same rule applies to more distant relationships (second-degree, third-degree, etc.) (20).

Based on the result from the KING-robust analysis, a principal component analysis was conducted using the PC-AiR package to estimate the ancestrally representative PCs (17). The genotyped data were filtered and pruned using the SNPRelate package in R, with a MAF threshold of 0.05, linkage disequilibrium threshold of √0.1, and a maximum sliding window of 10^6^ bp (21). Pairs of relatives with a kinship coefficient > 0.022 (2^11/2^) (fourth-degree relatives or closer) were defined as related; pairs with a kinship coefficient less than -0.022 were defined as unrelated (11).

A PC-Relate analysis was then used to generate an adjusted kinship matrix, to estimate the familial relatedness when adjusting for ancestral background (22). Unrelated participants and the first four ancestrally representative PCs were used to estimate such relationships. Any resulting kinship coefficients < 0.022 were sent to 0 to improve processing speed in the regression analyses (11).

***Adolescent Brain Cognitive Development (ABCD) Study***

Using the PCs, generated as above, a subset of participants who were ancestrally European were identified. Those with a first PC < -0.005 and a second PC < 0.002 were classified as ancestrally European, based on visual inspection of the plots generated (11). In this subset, there were 5,763 participants and 8,665,039 variants.

**Polygenic Score Generation**

PRS-CSx allows for the inclusion of multiple ancestries in the generation of a single polygenic score, which aims to improve cross-population prediction (23). For generation of the schizophrenia polygenic score, as separate GWAS summary statistics were available for four ancestry groups (Asian, African American, European, and Latino), all four were combined in the PGS generation, matched with the following 1000 Genomes reference panels (13):

- Asian ancestry summary statistics: East Asian (EAS) reference panel
- African American ancestry summary statistics: African (AFR) reference panel
- Latino ancestry summary statistics: Admixed American (AMR) reference panel
- European ancestry summary statistics: European (EUR) reference panel

As only European ancestry summary statistics were available for the other polygenic scores used (bipolar disorder, intelligence, and educational attainment), the EUR reference panel alone was used for these.

**Neuropsychological Test Descriptions**

| **Cognitive Test (Database)** | **Description** |
| --- | --- |
| Block design  (PEIC) | Participants were given a number of cube-shaped blocks that have faces of different colours: some are red, some are white, and some are split diagonally to be half red and half white. Participants were asked to use an increasing number of these blocks to replicate a pattern presented to them, which was first presented as a physical model made from the same type of blocks, and then presented as a two-dimensional picture (showing what the top surface of the blocks should look like) (24).  As the tests used different between research centres (WAIS-R or WAIS-III), scores reported have been converted to the percentage of the maximum possible score (participants’ score / maximum score for test). |
| Digit span  (PEIC) | Consists of two parts: digit forwards, in which participants must repeat back sequences of numbers presented auditorily in the same order that the digits were presented; and digits backwards, where participants must repeat back a sequence in reverse order (25).  As the tests used different between research centres (WAIS-R or WAIS-III), scores reported have been converted to the percentage of the maximum possible score (participants’ score / maximum score for test). |
| Rey Auditory Verbal Learning Test (RAVLT)  (PEIC) | Participants were presented auditorily with a list of 15 unrelated words over three learning trials, and asked to recall as many as they could after each trial. Participants’ performance on these trials constitutes their immediate recall score. After a 30-minute interval, participants were asked to recall as many words as they could from the first list of 15 words only – performance on this constitutes the delayed recall score. |
| Rey Auditory Verbal Learning Test (RAVLT)  (ABCD^®^ Study) | Participants were presented auditorily with a list of 15 unrelated words over five learning trials, and asked to recall as many as they could after each trial. Participants’ performance on these trials constitutes their immediate recall score. A new list of 15 words is then presented as a distractor; after recalling as many words as they can from this list, participants were then asked to recall as many words as they could from the first list (without repetition of the list beforehand) – performance on this constitutes the short-delay recall score. After a 30-minute interval, participants were asked to recall as many words as they could from the first list of 15 words only – performance on this constitutes the long-delay recall score (26). |
| NIH Toolbox Picture Vocabulary Test^®^ (TPVT)  (ABCD^®^ Study) | Participants heard a word accompanied by four images, one of which matched the word heard, and were asked to match the word to the image (26). |
| NIH Toolbox Oral Reading Recognition Test^®^ (TORRT)  (ABCD^®^ Study) | Participants were shown letters and words and asked to read them aloud, measuring their exposure to language materials and the cognitive skills involved in reading (26). |
| NIH Toolbox Pattern Comparison Processing Speed Test^®^ (TPCPST)  (ABCD^®^ Study) | Participants were shown two pictures and asked to determine whether they were the same or no. Scoring is based on how many trials participants can answer correctly within a given time frame (26). |
| NIH Toolbox List Sorting Working Memory Test^®^ (TLSWMT)  (ABCD^®^ Study) | Participants were presented with a series of pictures of animals or foods of different sizes, accompanied by the name of the image auditorily, and then asked to repeat back the items in order from smallest to largest. The test starts with images from just one of the two categories before combining the types of images shown (26). |
| NIH Toolbox Picture Sequence Memory Test^®^ (TPSMT)  (ABCD^®^ Study) | Participants were shown a series of 15 pictures depicting activities or events that occur within a particular setting (e.g. working on a farm) and asked to imitate the actions in the sequence as it was presented (26). |
| NIH Toolbox Flanker Task^®^ (TFT)  (ABCD^®^ Study) | Participants were shown a total of five arrows presented horizontally across the screen. The two outer arrows on each side all faced the same way, either left or right, and the middle arrow faced either the same direction as the others (congruent trials: 🡨🡨🡨🡨🡨) or the opposite direction (incongruent trials: 🡨🡨🡪🡨🡨). Participants were asked to indicate which direction, left or right, the arrow is pointing. Scoring is based on both speed and accuracy (26). |
| NIH Toolbox Dimensional Change Card Sort^®^ (TDCCS)  (ABCD^®^ Study) | Participants were presented with two objects at the bottom of the screen and a third in the middle of the screen, which they were asked to sort with one of the two objects at the bottom based on either colour or shape. In the first block of trials, the participant sorts the object on one of the dimensions (e.g. colour), on the second they sort the object with the other dimension (e.g. shape), and during the third block the dimension to be used alternates in pseudorandom order between the two. Scoring is based on accuracy and speed of the participants’ responses (26). |
| Cash Choice Task  (ABCD^®^ Study) | Children were asked “Let’s pretend that a kind person wanted to give you some money. Would you rather have $75 in three days, or $115 in three months”, to which they can respond with either monetary option, or “can’t decide”. The first option is considered to reflect a preference for immediate gratification and the second is considered to reflect a preference for delayed gratification (26). |
| Little Man Task  (ABCD^®^ Study) | A simplistic male figure is shown, holding a briefcase in either his left or right hand. The figure is shown either right-side-up or upside-down, and either facing the participant or facing away. On each trial, the child was asked to determine which hand, left or right, was holding the briefcase (26). |
| Matrix Reasoning  (ABCD^®^ Study) | Participants were shown a visuospatial array consisting of a series of stimuli which is incomplete alongside four options presented below, and asked to determine which of the four options completed the sequence (26). |
| *Abbreviations:* PEIC = Psychosis Endophenotypes International Consortium; ABCD = Adolescent Brain Cognitive Development; NIH = National Institutes of Health | |

**Cognitive Test Performance**

***PEIC***

Supplementary Table S2 displays the unadjusted average cognitive test performance. Supplementary Table S3 displays effect estimates for the differences in performance between groups, controlled for age. The control group performed significantly better, on average, than patients on all tests (ps < 1.23x10^-12^). Controls performed significantly better than relatives on block design and digit span (ps < 1.17x10^-9^), but there was no statistical difference on the RAVLT (ps > .0741). Relatives performed significantly better than patients on the RAVLT (ps < 2x10^-16^) and block design (p = .0166), but significantly worse (p = .0448) on digit span.

***ABCD***

Average age-corrected cognitive test scores for the ABCD Study sample are presented in Supplementary Table S4.

**Interaction and Subgroup Analyses in the PEIC Dataset**

Subgroup analysis were carried out despite the non-significant interaction effects of clinical group on the effect of PGS on cognitive performance due to the previous evidence of a difference in the strength of the effect between patients and control participants (27). Fixed effect regression analyses were carried out (as the kinship matrix, included as a random effect in the main analyses, was not needed as no participants in the control group or patient group were related to each other). The covariates included were age, sex, research site, and the first four ancestry PCs. Results from the subgroup analyses in the control and patient groups are presented in Supplementary Figure S1 (Supplementary Tables S17 and S18). Neither of the psychosis-related polygenic scores showed a significant effect on performance on any of the cognitive tests when the samples were restricted to the subgroups after correction for multiple comparisons. The effect of cognition-related PGSs were similar between the groups for all tests.

This appears to contradict the findings in Mallet et al’s (27) meta-analysis, which found that, once samples are separated into psychosis cases and controls, the negative effect of schizophrenia PGS on cognitive performance is seen in controls only. However, interpretation of the subgroup results should consider the non-equal sample sizes (smaller patient sample than control) and that the accompanying interaction analyses were non-significant. Further investigation with larger, equal samples is needed.

**(a)**

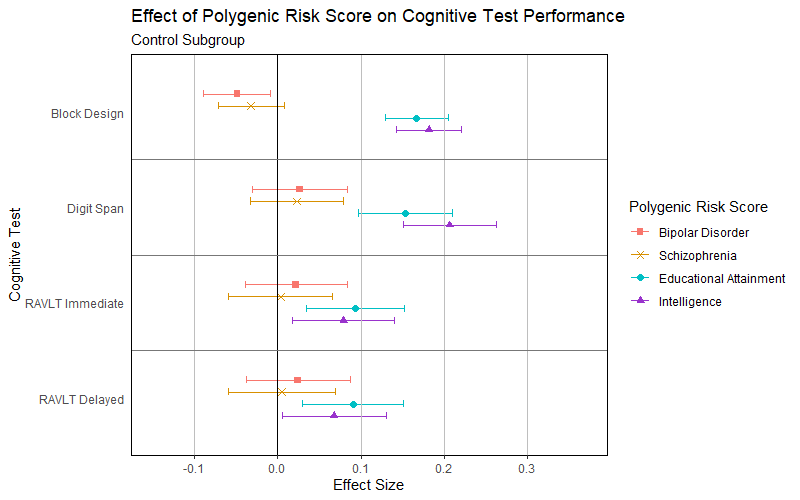

**(b)**

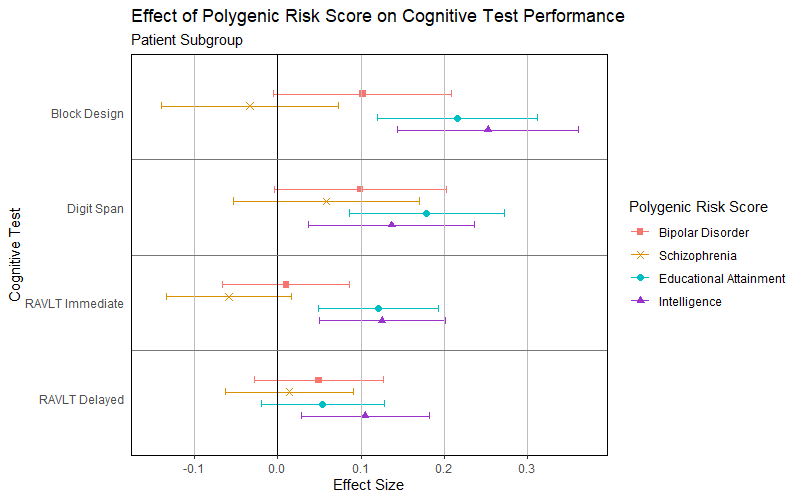

**Supplementary Figure S1. Effect of Polygenic Scores on Cognitive Test Performance in (a) the Control Group and (b) the Patient Group in the PEIC Sample**

Effect of psychosis-related and cognition-related polygenic scores on cognitive test performance in adults separately in (a) the control group (i.e. no diagnosis of psychosis or family relation to a patient with psychosis) and (b) the patient group (i.e. individuals with a diagnosis of psychosis), while controlling for the effect of age, gender, research site, and ancestry (the first four ancestry principal components). Scores have been standardised using the mean and standard deviation from the control group. Standardised values are given in Supplementary Table S5; non-standardised values are given in Supplementary Table S6.

*Abbreviations:* PEIC = Psychosis Endophenotype International Consortium; RAVLT = Rey Auditory Verbal Learning Task

**Table S1. Neurocognitive Test Domains for Bonferroni Correction to Analyses Using the ABCD Study**^®^ **Sample**

| **Cognitive Test (and Subtests)** | | **Cognitive Domain** |
| --- | --- | --- |
| NIH Toolbox Picture Vocabulary Test^®^ (TPVT) | | Language |
| NIH Toolbox Oral Reading Recognition Test^®^ (TORRT) | |  |
| NIH Toolbox Dimensional Change Card Sort^®^ (TDCCS) | | Executive functioning |
| NIH Toolbox Flanker Task^®^ (TFT) | |  |
| NIH Toolbox Pattern Comparison Processing Speed Test^®^ (TPCPST) | | Processing speed |
| NIH Toolbox List Sorting Working Memory Test^®^ (TLSWMT) | | Working memory |
| Little Man Task | % Correct | Visuospatial processing |
|  | % Incorrect |  |
| Matrix Reasoning | |  |
| NIH Toolbox Picture Sequence Memory Test^®^ (TPSMT) | | Episodic memory |
| Rey Auditory Verbal Learning Task (RAVLT) | Immediate Recall |  |
|  | Short Delayed Recall |  |
|  | Long Delayed Recall |  |
| Cash Choice Task | | Motivation/impulsivity |
| *Abbreviations:* ABCD = Adolescent Brain Cognitive Development | | |

**Predicting Clinical Group Status from Cognition-Related PGS Using a Support Vector Machine Algorithm**

***Methods***

To predict clinical groups, a binary classification Support Vector Machine algorithm (SVM) was employed. SVM is a supervised learning algorithm that can handle non-linear relationships within high-dimensional data (28) and it has been used with success in predicting clinical groups from genetic data (29) and other biomarkers (30). The analysis was carried out using Scikit-learn 1.3.2. and SciPy 1.11.4. packages of Python 3.9 (31).

For the PEIC data, binary classifications were performed for patient-control, patient-relative and relative-control comparisons. The predictors in the model were: age, sex, testing location, four ancestry PCs and intelligence or educational attainment PGS. For the ABCD data, the comparisons were between those who experienced psychosis-like symptoms at baseline and those who did not, those who experienced distressing psychosis-like symptoms and those who did not, and those who experienced significantly distressing psychosis-like symptoms and those who did not. The predictors were: age, sex, eight ancestry PCs and intelligence or educational attainment PGS.

As the analyses wanted to follow the regression as closely as possible, the pre-processing steps and scaling operations were the same. However, all participants with missing data were excluded instead of the missing variables being imputed, as within the nested cross-validation procedure this would have risked data leakage. In addition, the testing location variable in the PEIC data was dummy coded.

To mirror the regression’s random effect from the adjusted kinship matrix, this variable was incorporated in the kernel operations during the SVM classification. The SVM models used a custom kernel operation, that summed together the adjusted kinship matrix with the Gram matrix of a regular RBF kernel. The weight of the kinship matrix and the RBF kernel’s Gram matrix added up to 1 (Supplementary Equation S1). The weight component was used as a hyperparameter w and as for any other hyperparameter, its value was determined following a parameter search.

*R* = *w* · *G* + (1 *- w*) *· K*

*R:* the resulting pairwise similarity matrix of the data

*w:* weight (coefficient) that scales the Gram matrix.

*G:* Gram matrix resulting from the regular RBF kernel transformation.

*K:* kinship matrix.

*(1−w):* scales the kinship matrix

**Supplementary Equation S1. Custom kernel operation in the SVM model**

The other hyperparameters for the SVM models were C, γ and W. C and γ are part of every RBF kernel SVM model as the regularisation parameter and the radius of influence for training samples, respectively. The hyperparameter W was defined as the weight of the clinical group membership during training phase of the SVM pipeline. This was done to offset the imbalance between the clinical group memberships.

The analysis pipeline took the form of a nested cross-validation, to reduce biases in the hyperparameter tuning. The general structure of this followed the methods of Bracher-Smith et al. (32) with having 10 inner cross-validation folds to determine the best hyperparameters and 10 outer cross-validation folds to evaluate the performance of the SVM model with the selected hyperparameters. Inner cross-validation was only run on the training set, to avoid data leakage.

For each round of outer cross-validation, 100 hyperparameters combinations were drawn using random parameter search (33). C and γ were sampled from the same distributions as in Bracher-Smith et al. (32). As there was no preconceived constraint for the value of w, it was sampled from a uniform distribution between 10-6 and 1-10-6. W was sampled from a uniform distribution between 1 and 4. The probability density functions of the hyperparameter sampling are depicted in Supplementary Figure S2. Each combination of hyperparameters was tested on 10 inner cross-validation loops and the combination with the highest F1 score was selected as the best. F1 scores were used, as this metric balances the classification errors between false negatives and false positives. After selecting the best hyperparameters, the SVM model was fit on the training dataset. Then, classification performance was evaluated on the unseen test set using measures of accuracy, precision, recall, F1 scores and the areas under the ROC curve (AUROC).

The fitted SVM model was also subjected to a permutation feature importance (PFI) analysis (34), to determine the relative importance of each predictor. This meant that for each predictor in the test set, the values were randomly shuffled between the participants and the SVM model was tested again on this novel data. The decrease in AUROC indicated the importance of the predictor. These values were normalized across predictors, to get a relative measure of importance across the model.

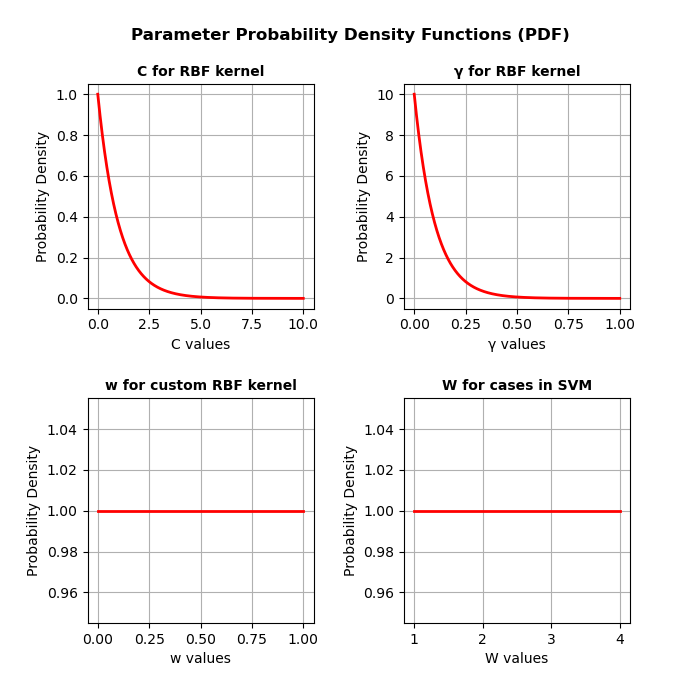

**Supplementary Figure S2. Parameter Probability Density Functions for hyperparameter tuning using random search**

The probability densities of the C, γ, w (Gram matrix weight) and W (case class weight) hyperparameters, from which the 100 iterations of hyperparameters were sampled randomly. The PDF for C and γ was adapted from Bracher-Smith et al. 2022 and the uniform distribution of w and W values were employed as exploratory measures.

***Results***

In the PEIC dataset, there were 321 missing age values. After excluding these participants, the analysis was performed on a sample with N = 3,650.

The results of the random hyperparameter search for case-control classification for both SVM models are depicted in Supplementary Figure S3. Generally, higher weighting of the demographic and PGS predictors compared to the kinship matrix yielded a better model performance. This is accompanied by low γ and C parameters. The weight of cases (patients or relatives) compared to controls generally took higher values in the successful classification tasks and low values in the relative-control classification tasks.

The classification performance metrics for both SVM models and classification tasks are depicted in Table S19. The discrimination in the patient-control and patient-relative cases seems robust with high AUROC and moderately high accuracy scores. This corroborates the regression analysis and goes beyond its predictive value in the patient-relative classification case. However, in the relative-control case, there is a large imbalance between positive and negative class predictions, as accuracy and precision scores are always lower than the corresponding recall scores. Due to the almost chance accuracy, these classification performances were determined as not successful. To illustrate the sensitivity-specificity trade-off, Receiver Operating Characteristics (ROC) curves are plotted for each of the 10 outer cross-validation folds for each analysis in Supplementary Figure S4.

The permutation feature analysis revealed the predictors with the highest relative importance in each model. While the regression analysis assessed the separate predictive value of the PGS, this analysis served as a way of comparing the contribution of the PGS to the rest of the model predictors in a model-agnostic fashion. Across the analyses, PGS as a predictor never took up a considerable (M > 0.1) amount of importance. Instead, the most important contributor seemed to be age, and in other cases, ancestry and research sites. These results are illustrated in Supplementary Figure S5. The importance of age and research sites in the model performance are not surprising, given the age imbalance in the data and for some sites providing only patient or only control data.

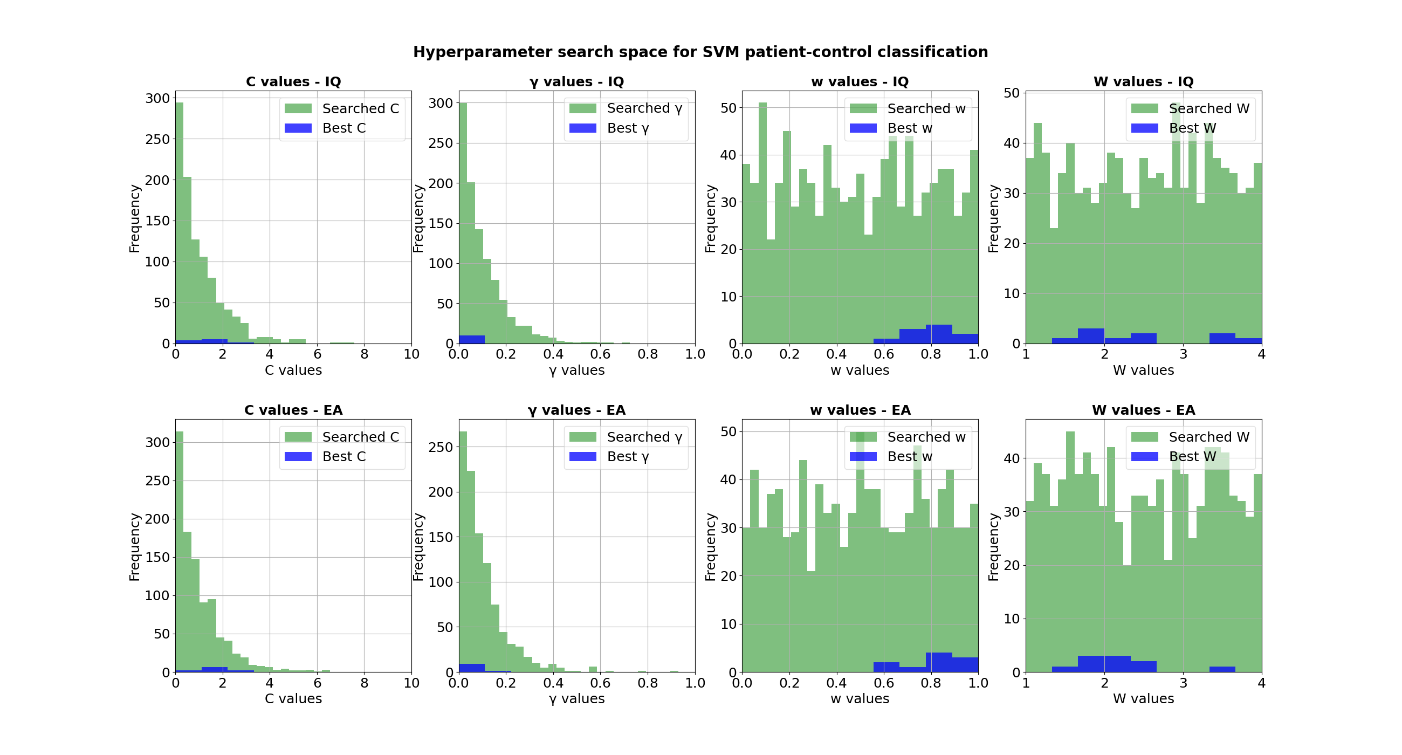

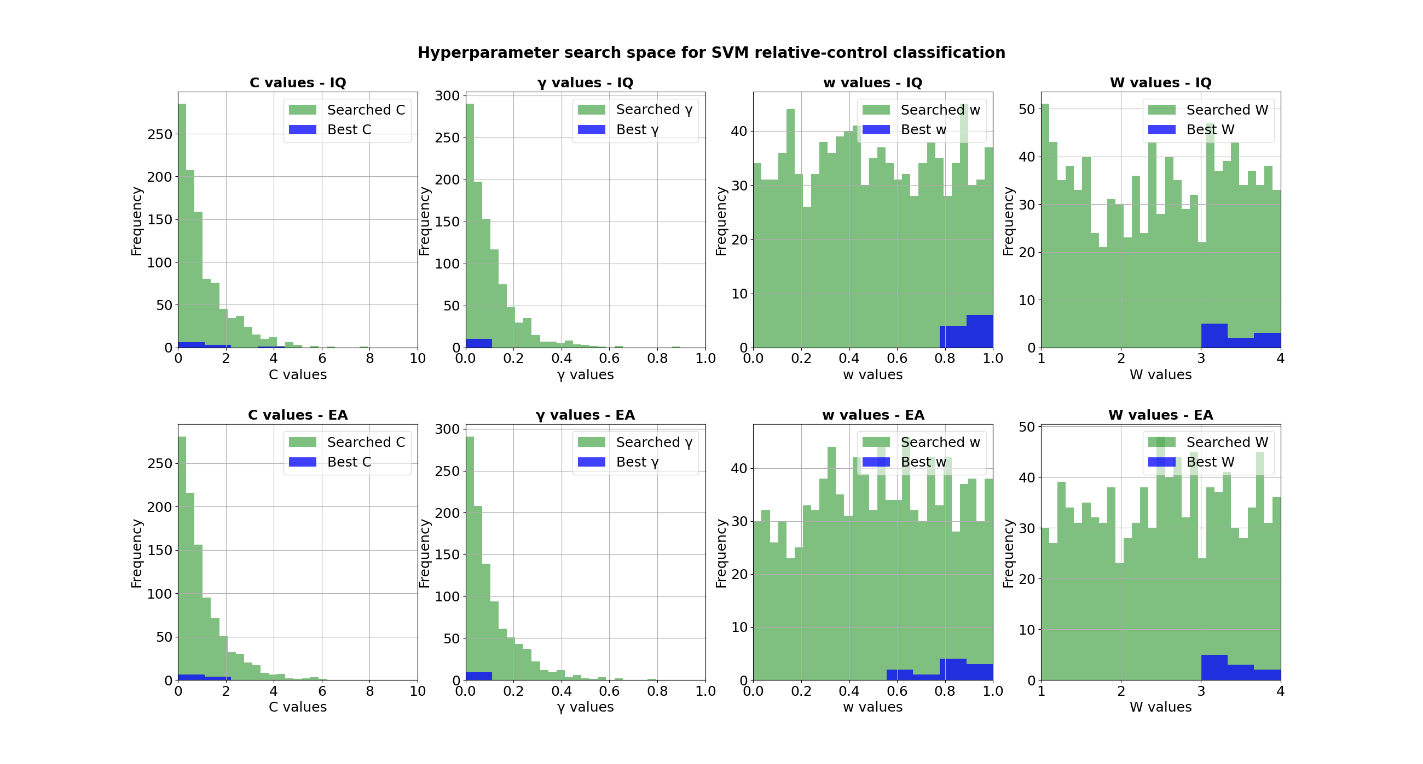

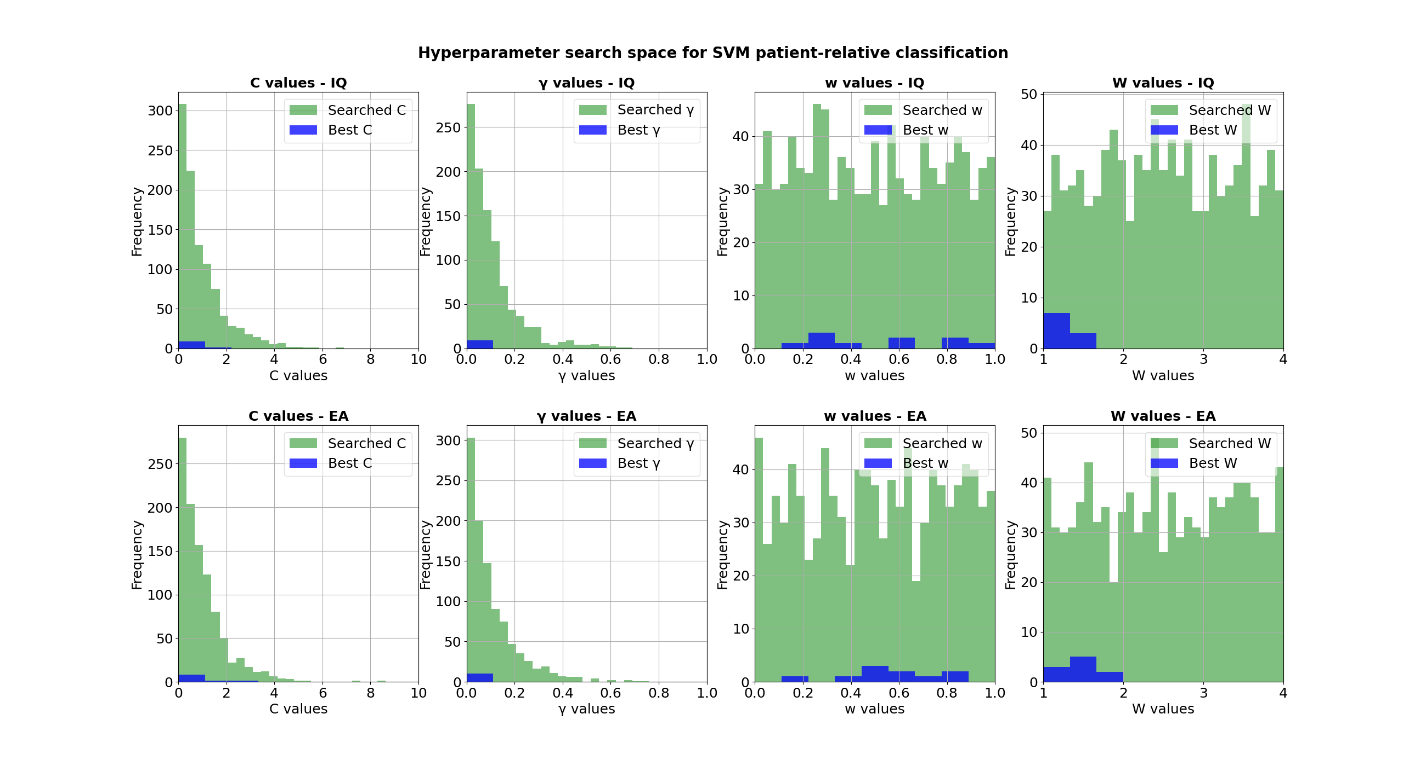

**Supplementary Figure S3. Hyperparameters searched and used during the clinical group binary classifications**

Binned C, γ, w (Gram matrix weight) and W (case class weight) hyperparameter values that were used in each iteration of the random parameter search (green) and the ones that were selected as best parameters (blue). Best parameters were selected based on highest F1 scores on the 10 inner folds. The histograms display hyperparameter values across all 10 outer folds. The top row shows results of the model using intelligence PGS, the bottom row shows the model with the educational attainment PGS.

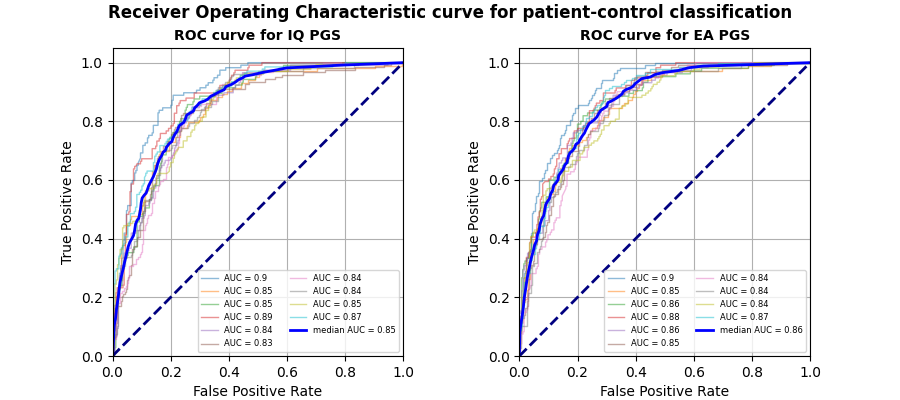

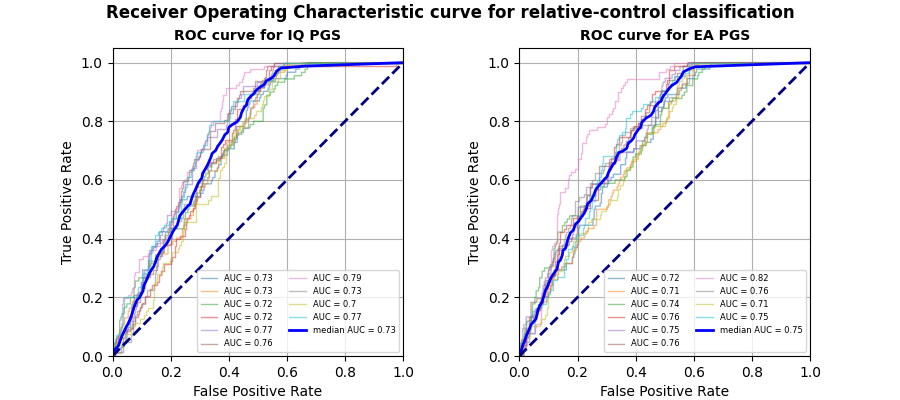

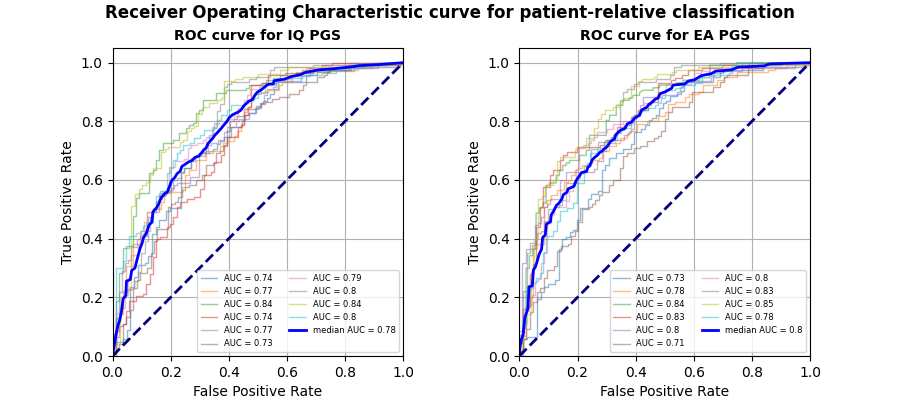

**Supplementary Figure S4. ROC curves for clinical group classification using SVM models with IQ and EA predictors**

ROC curves are displayed for patient-control, relative-control and patient-relative binary classification tasks. ROC curves plot the false positive rate to the true positive rate, to display the sensitivity-specificity trade-off of the classifier. Each figure depicts the ROC curves for each of the 10 outer folds of cross-validation. Their corresponding measures for the area under the curve (AUC) are reported in the figure legend. The blue line represents the mean of such curves, it is reported with the median AUC. The dashed line represents a classifier at chance level.

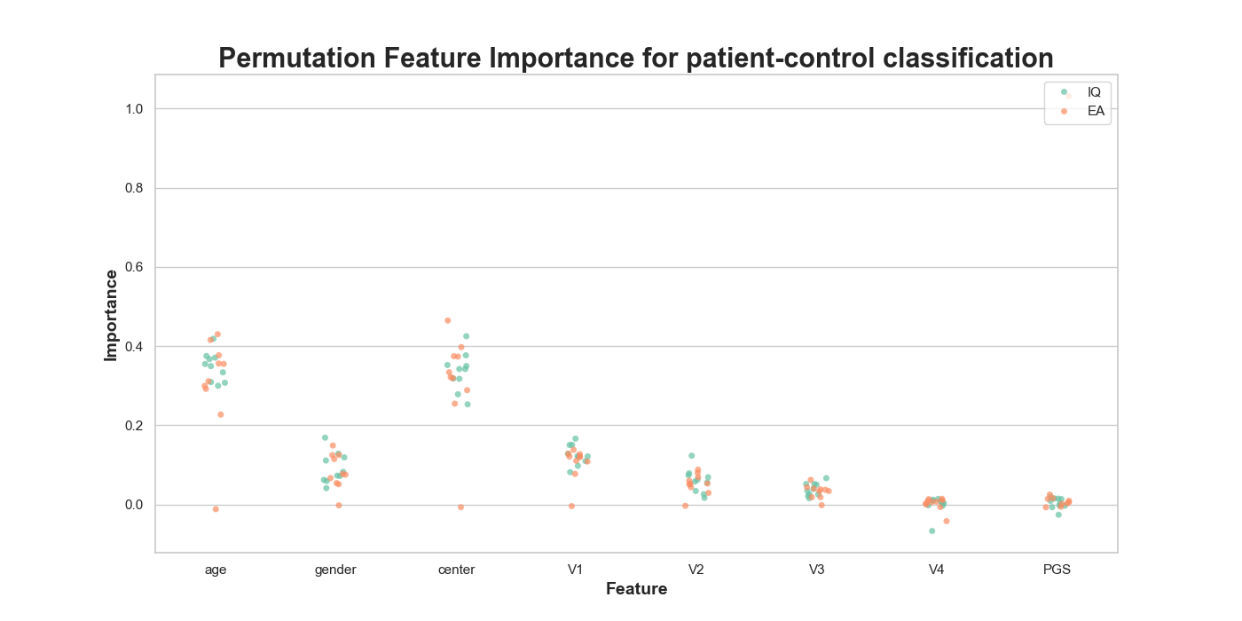

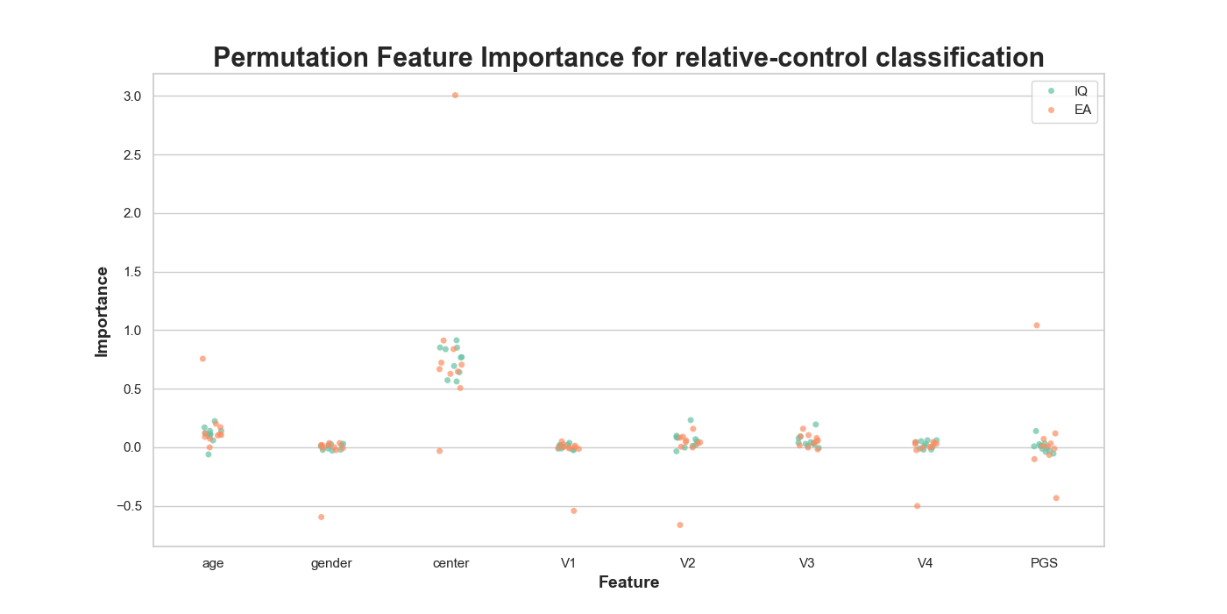

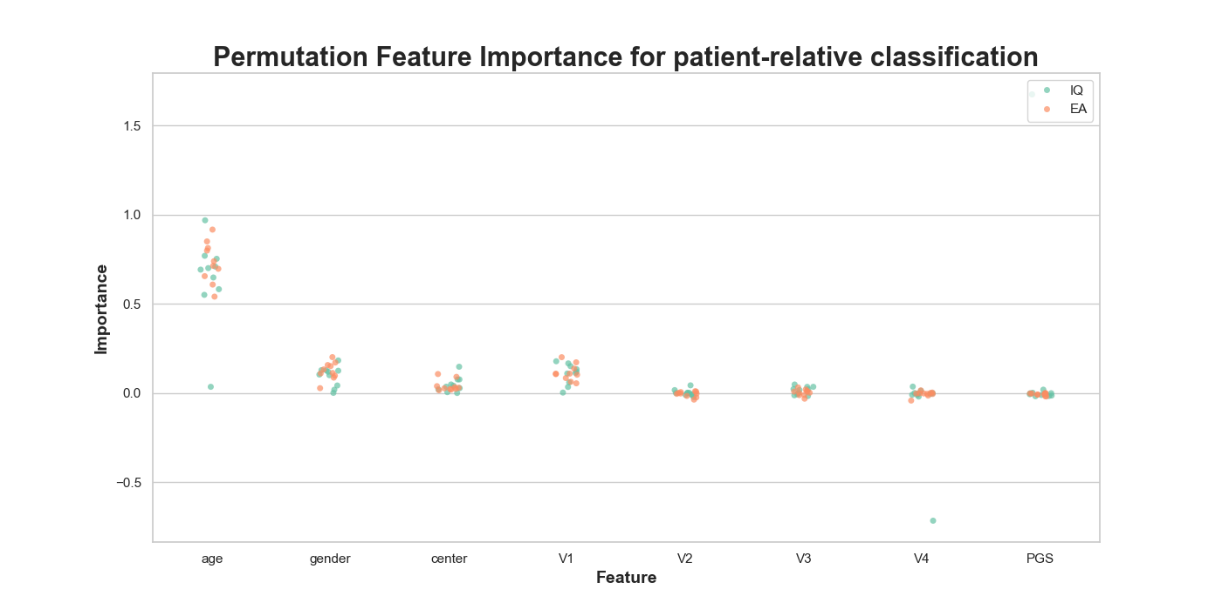

**Supplementary Figure S5. PFI scores for clinical group classification using SVM models with IQ and EA predictors**

Relative feature importance scores are displayed for each model predictor in each clinical group classification task. Each clinical group classification is on a separate graph. Model predictors are shown on the x axis, with PGS being IQ and EA in corresponding models. Features V1 to V4 are the ancestry principal components used in the model Feature importance scores are indicated on the y axis. They show relative importance, as they are normalized to 1 across features. Scores for IQ PGS models are green, scores for EA PGS models are orange.

In the ABCD dataset no further participants were excluded due to missing data. The results of the hyperparameter random search are displayed in Supplementary Figure S6. Generally, the models performed best with low C and γ parameters and with a generally high Gram matrix weight w. The weight of the case group W ranged across all potential values. However, as the model performance was generally poor, these tendencies convey little meaning.

The classification performance of the SVM models for each psychosis-like symptom group is depicted in Table S20. The ROC curves indicating the sensitivity-specificity trade-off are in Supplementary Figure S7. The discrimination performance and the classification accuracy, as indicated by the AUROC and accuracy values were near chance level for each model and classification task, suggesting a poor model performance. However, the high recall scores indicate that what the models excelled at was the classification of true positives, that is, classifying specifically case category data.

While these performance metrics suggest that the SVM model was unable to validate the results of the main regression analysis, there are also a few model and dataset specifications that could be the reason behind it. Firstly, optimising for F1 scores happens at the cost of accuracy, so potentially an accuracy-optimised model would have been able to have a better predictive value. Secondly, the imbalance in the distressing psychotic symptom-control and significantly distressing psychotic symptom-control comparisons was the highest amongst all comparisons and datasets. Therefore, a W parameter adjusted to these imbalances (e.g. searched between 1 and 10) might have resulted in a better model performance. Regarding the sample itself, it is also important to note that psychosis clinical groups have no relevancy at this young age. Rather, comparisons were made between categories of psychosis-prone symptoms that are experienced even amongst the non-clinical population. Nevertheless, it would be interesting to test whether the SVM model is able to perform classification on the follow-up ABCD data, when psychosis clinical groups become more clearly separated.

The results of the PFI analysis are displayed in Supplementary Figure S8. Again, due to the generally poor model performance, these tendencies do not convey too much relevant information as to which predictors might be the most important for a good model.

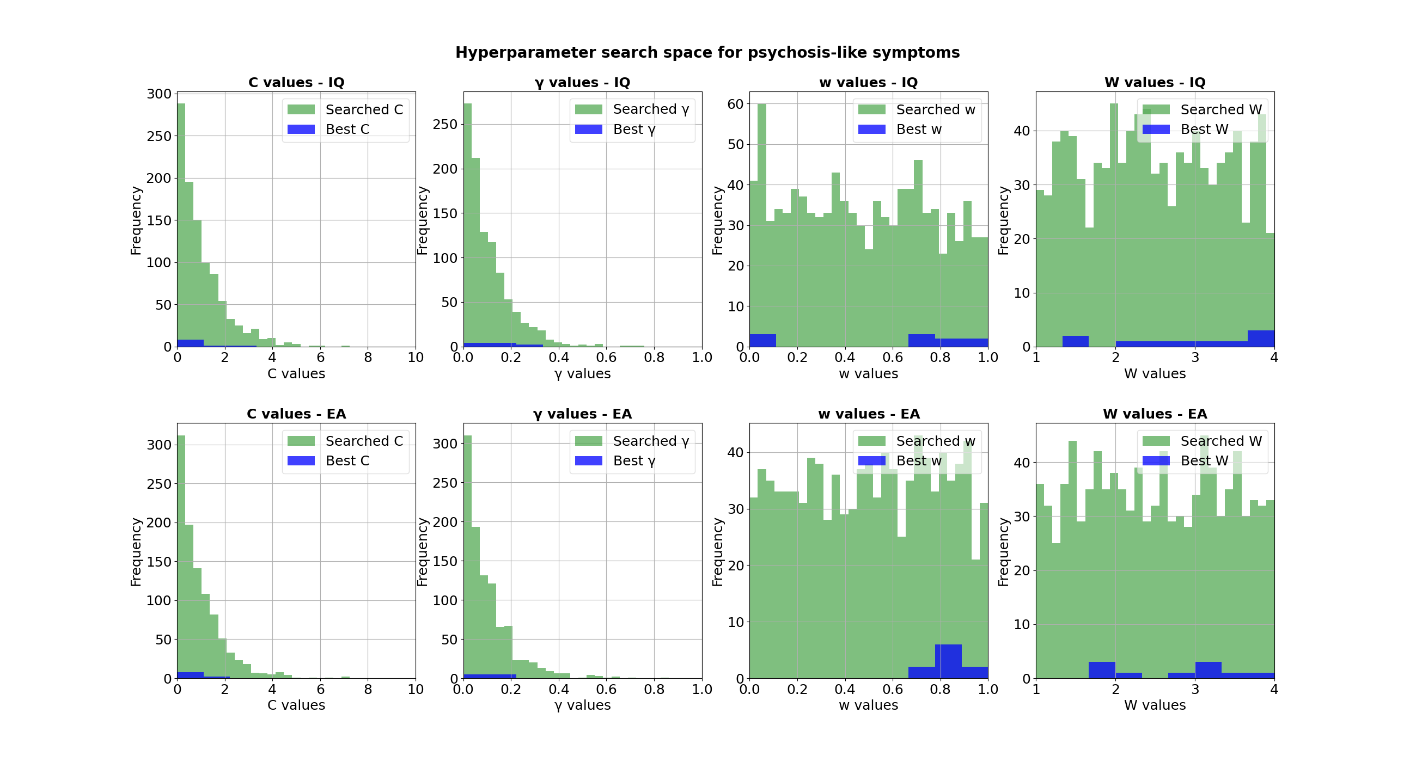

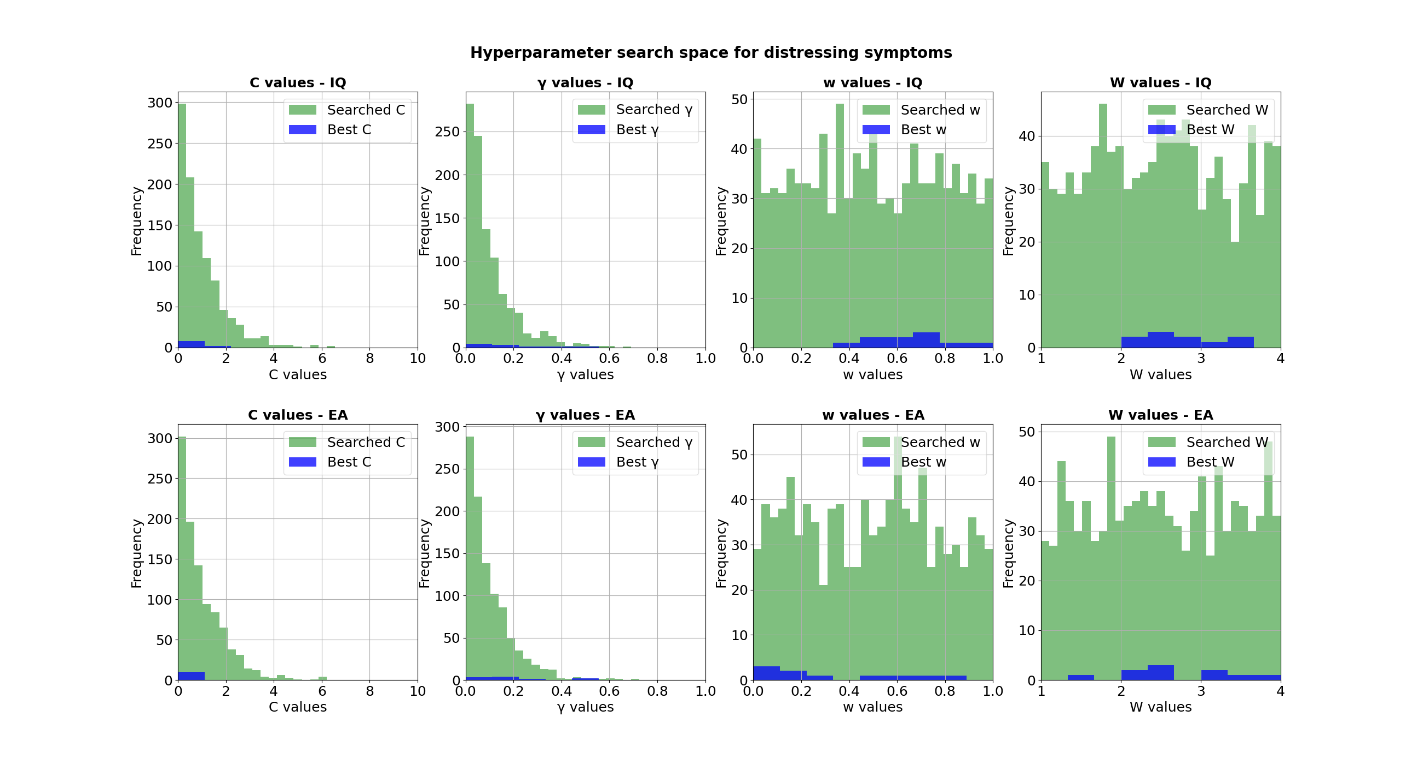

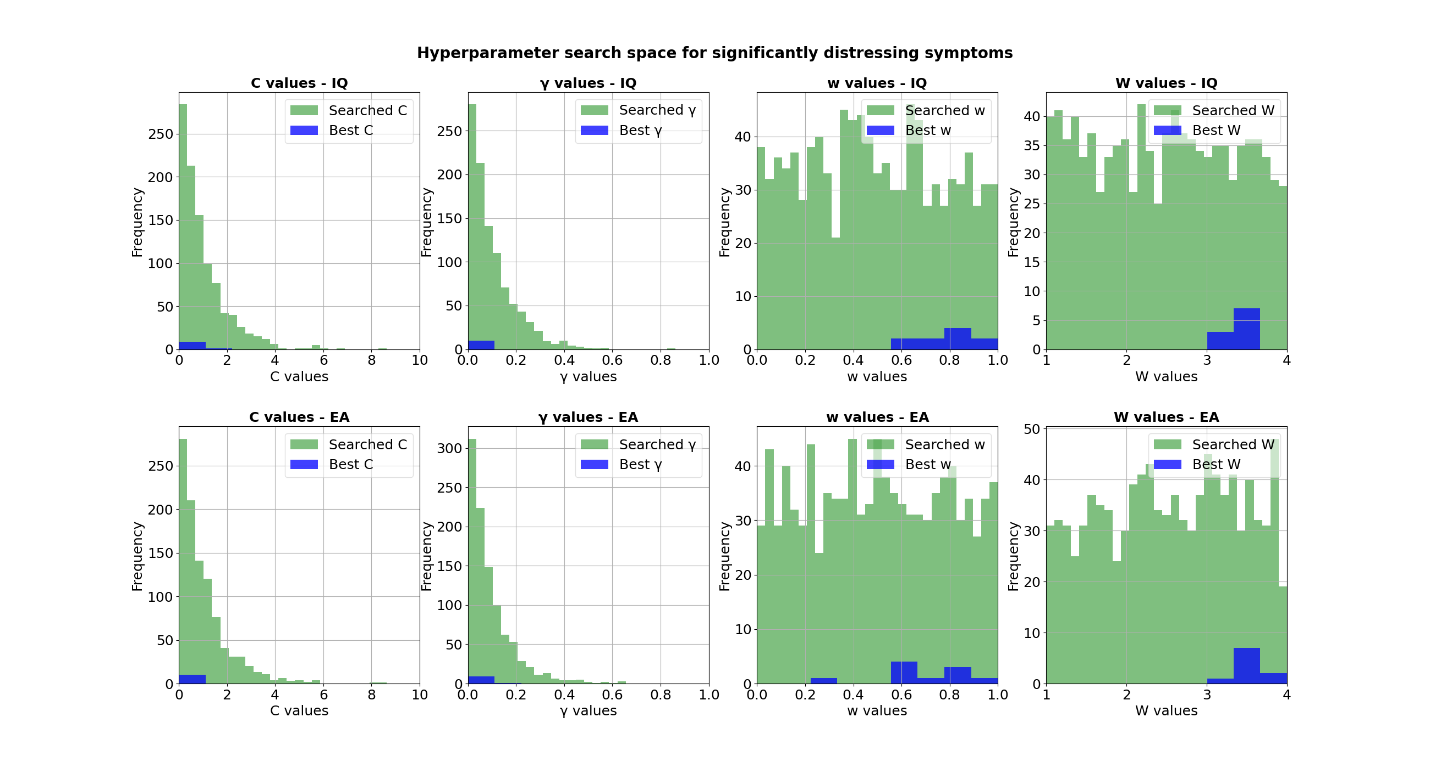

**Supplementary Figure S6. Hyperparameters searched and used during the clinical group binary classifications**

Binned C, γ, w (Gram matrix weight) and W (case class weight) hyperparameter values that were used in each iteration of the random parameter search (green) and the ones that were selected as best parameters (blue). Best parameters were selected based on highest F1 scores on the 10 inner folds. The histograms display hyperparameter values across all 10 outer folds. The top row shows results of the model using intelligence PGS, the bottom row shows the model with the educational attainment PGS.

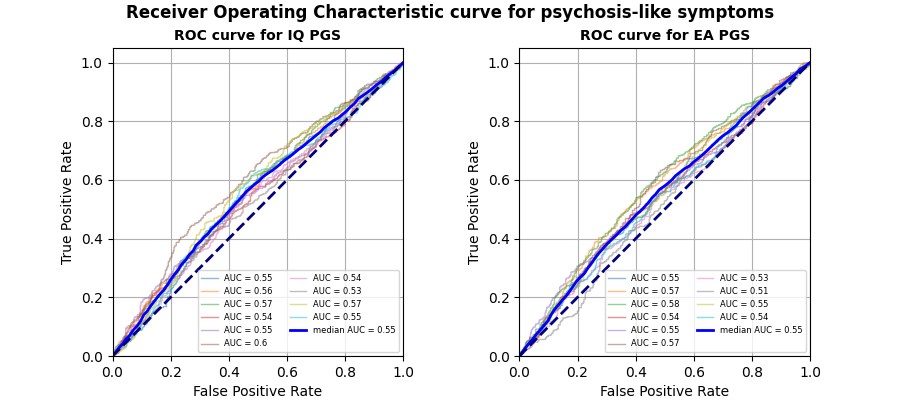

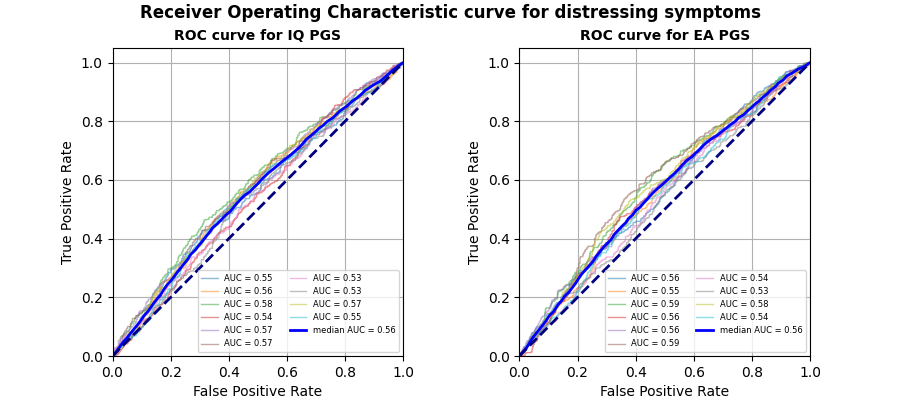

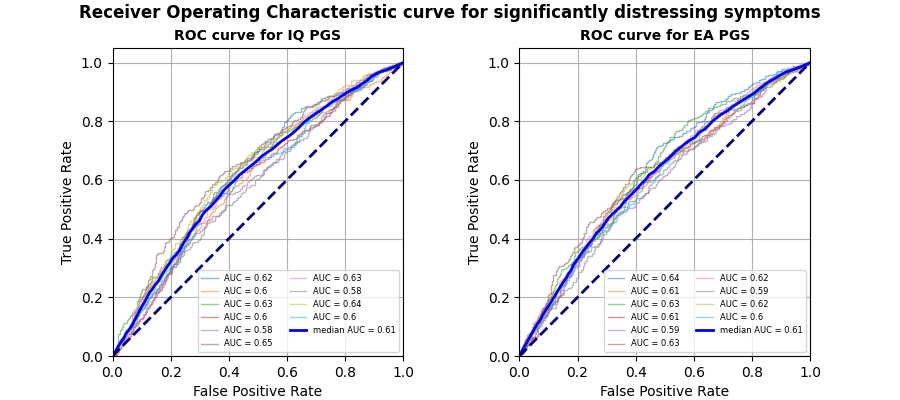

**Supplementary Figure S7. ROC curves for clinical group classification using SVM models with IQ and EA predictors**

ROC curves are displayed for binary classification tasks between participants who have and do not have psychosis-like symptoms, between participants who have and do not have distressing psychosis-like symptoms and between participants who have and do not have significantly distressing psychosis-like symptoms. ROC curves plot the false positive rate to the true positive rate, to display the sensitivity-specificity trade-off of the classifier. Each figure depicts the ROC curves for each of the 10 outer folds of cross-validation. Their corresponding measures for the area under the curve (AUC) are reported in the figure legend. The blue line represents the mean of such curves, it is reported with the median AUC. The dashed line represents a classifier at chance level.

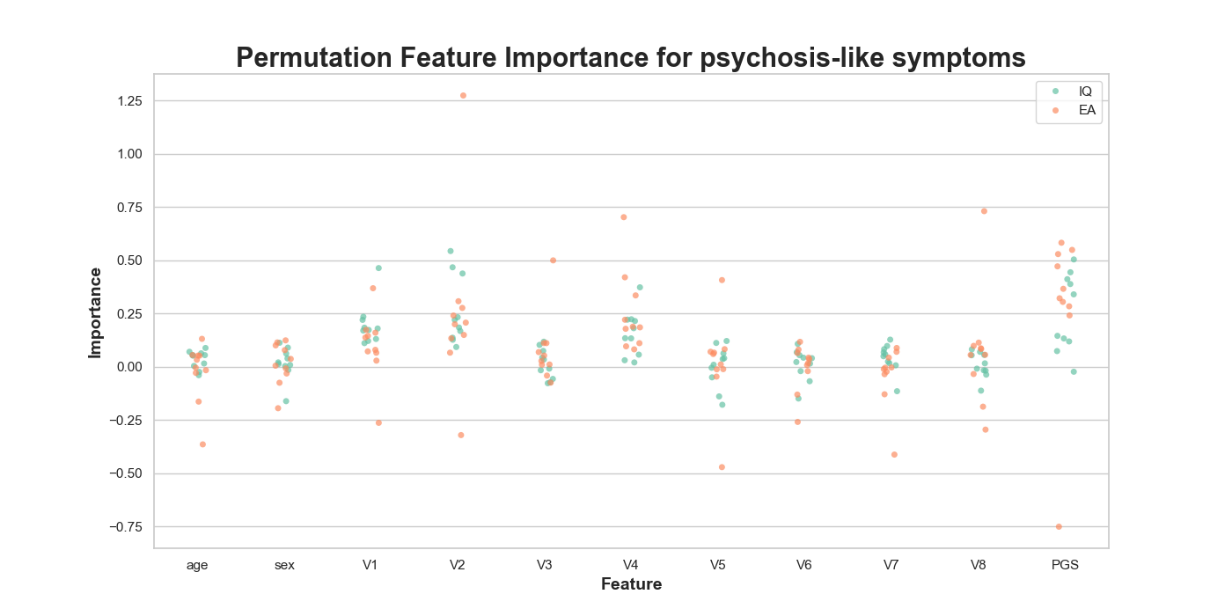

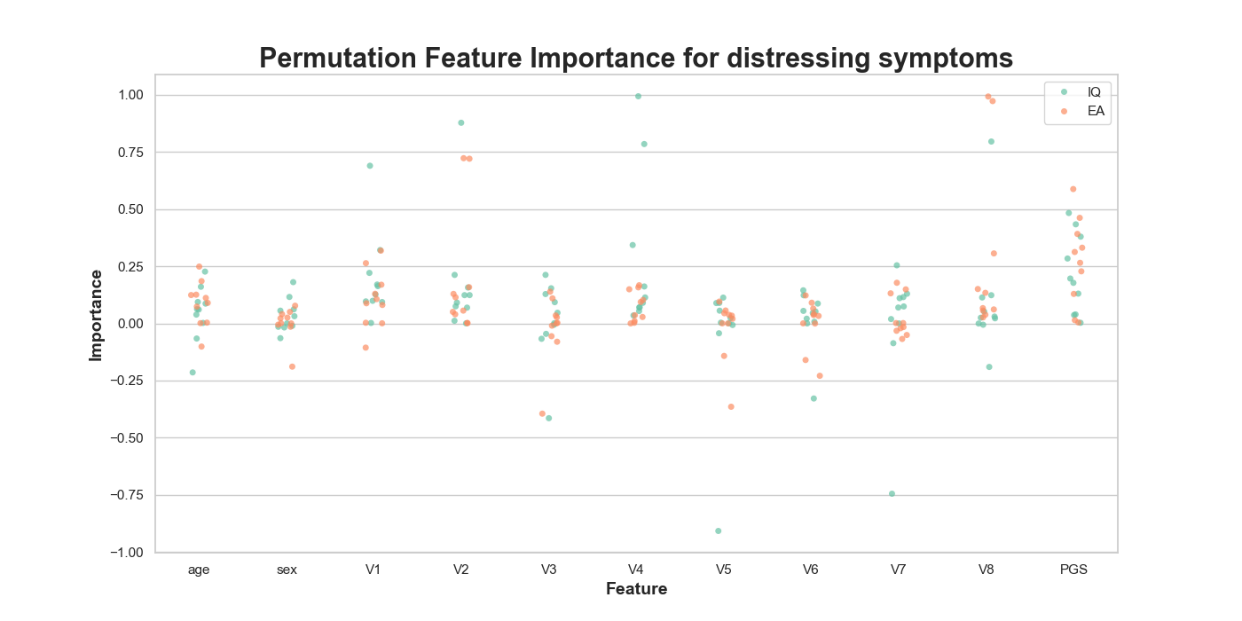

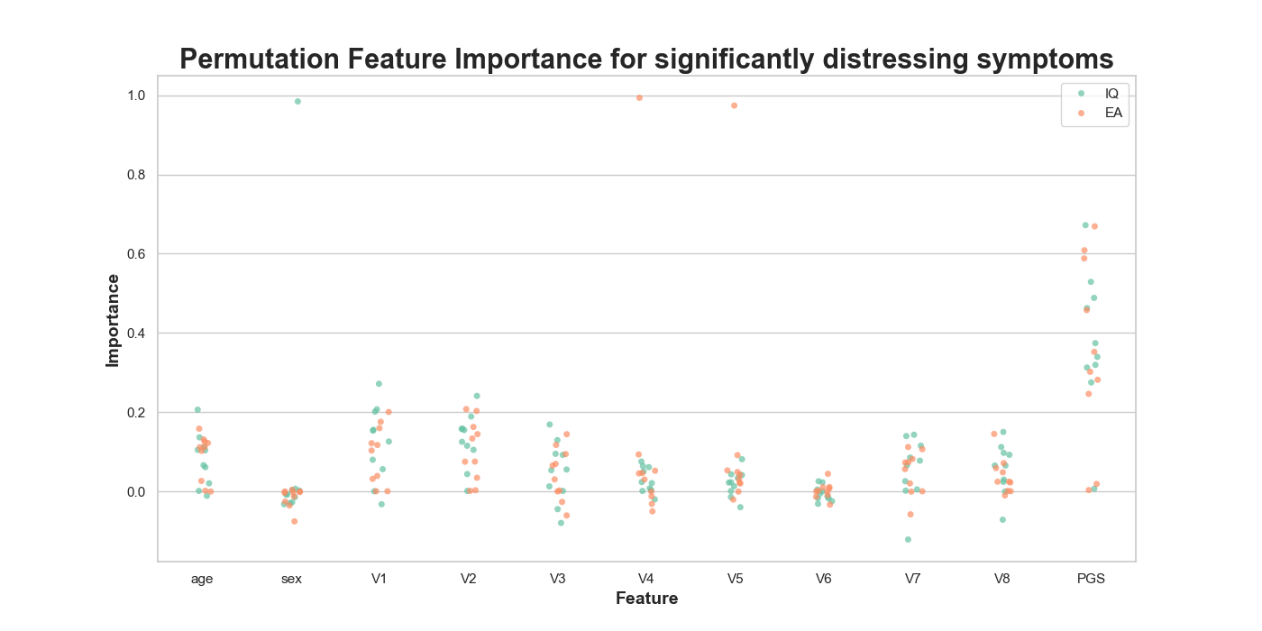

**Supplementary Figure S8. PFI scores for clinical group classification using SVM models with IQ and EA predictors**

Relative feature importance scores are displayed for each model predictor in each clinical group classification task. Each clinical group classification is on a separate graph. Model predictors are shown on the x axis, with PGS being IQ and EA in corresponding models. Features V1 to V8 are the ancestry principal components used in the model Feature importance scores are indicated on the y axis. They show relative importance, as they are normalized to 1 across features. Scores for IQ PGS models are green, scores for EA PGS models are orange.

**Supplementary Material References**

1.	Wellcome Trust Case Control Consortium. Genome-wide association study of 14,000 cases of seven common diseases and 3,000 shared controls. Nature. 2007;447(7145):661-78.

2.	Marchini J, Howie B, Myers S, McVean G, Donnelly P. A new multipoint method for genome-wide association studies by imputation of genotypes. Nat Genet. 2007;39(7):906-13.

3.	Wigginton JE, Abecasis GR. PEDSTATS: descriptive statistics, graphics and quality assessment for gene mapping data. Bioinformatics. 2005;21(16):3445-7.

4.	Morris JA, Randall JC, Maller JB, Barrett JC. Evoker: a visualization tool for genotype intensity data. Bioinformatics. 2010;26(14):1786-7.

5.	Speed D, Cai N, Johnson MR, Nejentsev S, Balding DJ. Reevaluation of SNP heritability in complex human traits. Nat Genet. 2017;49(7):986-92.

6.	Purcell S, Neale B, Todd-Brown K, Thomas L, Ferreira MA, Bender D, et al. PLINK: a tool set for whole-genome association and population-based linkage analyses. Am J Hum Genet. 2007;81(3):559-75.

7.	Bramon E, Pirinen M, Strange A, Lin K, Freeman C, Bellenguez C, et al. A genome-wide association analysis of a broad psychosis phenotype identifies three loci for further investigation. Biol Psychiatry. 2014;75(5):386-97.

8.	Durbin R. Efficient haplotype matching and storage using the positional Burrows-Wheeler transform (PBWT). Bioinformatics. 2014;30(9):1266-72.

9.	Loh PR, Danecek P, Palamara PF, Fuchsberger C, Y AR, H KF, et al. Reference-based phasing using the Haplotype Reference Consortium panel. Nat Genet. 2016;48(11):1443-8.

10.	Baurley JW, Edlund CK, Pardamean CI, Conti DV, Bergen AW. Smokescreen: a targeted genotyping array for addiction research. BMC Genomics. 2016;17(1):145.

11.	Wang B, Giannakopoulou O, Austin-Zimmerman I, Irizar H, Harju-Seppanen J, Zartaloudi E, et al. Adolescent Verbal Memory as a Psychosis Endophenotype: A Genome-Wide Association Study in an Ancestrally Diverse Sample. Genes (Basel). 2022;13(1).

12.	Lam M, Awasthi S, Watson HJ, Goldstein J, Panagiotaropoulou G, Trubetskoy V, et al. RICOPILI: Rapid Imputation for COnsortias PIpeLIne. Bioinformatics. 2020;36(3):930-3.

13.	Auton A, Abecasis GR, Altshuler DM, Durbin RM, Abecasis GR, Bentley DR, et al. A global reference for human genetic variation. Nature. 2015;526(7571):68-74.

14.	McCarthy S, Das S, Kretzschmar W, Delaneau O, Wood AR, Teumer A, et al. A reference panel of 64,976 haplotypes for genotype imputation. Nat Genet. 2016;48(10):1279-83.

15.	Loh PR, Palamara PF, Price AL. Fast and accurate long-range phasing in a UK Biobank cohort. Nat Genet. 2016;48(7):811-6.

16.	Das S, Forer L, Schönherr S, Sidore C, Locke AE, Kwong A, et al. Next-generation genotype imputation service and methods. Nat Genet. 2016;48(10):1284-7.

17.	Li H. A statistical framework for SNP calling, mutation discovery, association mapping and population genetical parameter estimation from sequencing data. Bioinformatics. 2011;27(21):2987-93.

18.	Gogarten SM, Sofer T, Chen H, Yu C, Brody JA, Thornton TA, et al. Genetic association testing using the GENESIS R/Bioconductor package. Bioinformatics. 2019;35(24):5346-8.

19.	R Core Team. R: A language and environment for statistical computing. R Foundation for Statistical Computing, Vienna, Austria. 2020 [Available from: <https://www.R-project.org/>.

20.	Manichaikul A, Mychaleckyj JC, Rich SS, Daly K, Sale M, Chen WM. Robust relationship inference in genome-wide association studies. Bioinformatics. 2010;26(22):2867-73.

21.	Zheng X, Levine D, Shen J, Gogarten SM, Laurie C, Weir BS. A high-performance computing toolset for relatedness and principal component analysis of SNP data. Bioinformatics. 2012;28(24):3326-8.

22.	Conomos MP, Reiner AP, Weir BS, Thornton TA. Model-free Estimation of Recent Genetic Relatedness. Am J Hum Genet. 2016;98(1):127-48.

23.	Ruan Y, Lin Y-F, Feng Y-CA, Chen C-Y, Lam M, Guo Z, et al. Improving polygenic prediction in ancestrally diverse populations. Nature Genetics. 2022;54(5):573-80.

24.	Soto T, Kraper C. Block Design Subtest. In: Volkmar FR, editor. Encyclopedia of Autism Spectrum Disorders. New York, NY: Springer New York; 2013. p. 464-5.

25.	Wambach D, Lamar M, Swenson R, Penney DL, Kaplan E, Libon DJ. Digit Span. In: Kreutzer JS, DeLuca J, Caplan B, editors. Encyclopedia of Clinical Neuropsychology. New York, NY: Springer New York; 2011. p. 844-9.

26.	Luciana M, Bjork JM, Nagel BJ, Barch DM, Gonzalez R, Nixon SJ, Banich MT. Adolescent neurocognitive development and impacts of substance use: Overview of the adolescent brain cognitive development (ABCD) baseline neurocognition battery. Dev Cogn Neurosci. 2018;32:67-79.

27.	Mallet J, Le Strat Y, Dubertret C, Gorwood P. Polygenic Risk Scores Shed Light on the Relationship between Schizophrenia and Cognitive Functioning: Review and Meta-Analysis. J Clin Med. 2020;9(2).

28.	Cortes C, Vapnik V. Support-vector networks. Machine Learning. 1995;20(3):273-97.

29.	Bracher-Smith M, Crawford K, Escott-Price V. Machine learning for genetic prediction of psychiatric disorders: a systematic review. Mol Psychiatry. 2021;26(1):70-9.

30.	Suri G-S, Kaur G, Moein S. Machine Learning in Detecting Schizophrenia: An Overview. Intelligent Automation \& Soft Computing. 2021;27(3):723--35.

31.	Pedregosa F, Varoquaux G, Gramfort A, Michel V, Thirion B, Grisel O, et al. Scikit-learn: Machine Learning in Python. Journal of Machine Learning Research. 2011;12(85):2825-30.

32.	Bracher-Smith M, Rees E, Menzies G, Walters JTR, O'Donovan MC, Owen MJ, et al. Machine learning for prediction of schizophrenia using genetic and demographic factors in the UK biobank. Schizophr Res. 2022;246:156-64.

33.	Bergstra J, Bengio Y. Random Search for Hyper-Parameter Optimization. Journal of Machine Learning Research. 2012;13(10):281-305.

34.	Altmann A, Toloşi L, Sander O, Lengauer T. Permutation importance: a corrected feature importance measure. Bioinformatics. 2010;26(10):1340-7.
